# Altered striatal functional gradients in obsessive-compulsive disorder

**DOI:** 10.1101/2025.07.15.25331605

**Authors:** Lachlan Webb, Luke Hearne, Ye E. Tian, Andrew Zalesky, Conor Robinson, Caitlin V. Hall, Saurabh Sonkusare, Bjorn Burgher, Michael Breakspear, Garance M. Meyer, Andreas Horn, Sebastien Naze, Philip Mosley, Luca Cocchi

## Abstract

**Background:** Obsessive-compulsive disorder (OCD) is associated with functional alterations in how the striatum interacts with the rest of the brain. However, the characterization of these changes in OCD is incomplete. Mapping functional striatal gradients provides a new opportunity to fill this knowledge gap. These gradients provide a spatial representation of continuous changes in whole-brain connectivity within striatal regions. Thus, OCD-related differences in striatal gradients imply changes in the functional organisation of striatal connections.

**Methods:** We calculated spatial striatal gradients linked to whole brain activity in 52 people with OCD and 45 controls. Gradients were computed with individuals at rest and when they underwent a threat-safety reversal task. Using a longitudinal dataset of 47 people with OCD, we investigated possible associations between changes in striatal gradient topology and fluctuations in symptom severity.

**Results:** Results showed group differences in the main gradient topology at rest, specifically in striatal regions overlapping with the putamen and caudate. Individuals showing a reduction in symptoms over time tended to change their gradient topology in favour of the control participants’ average topology. Finally, gradients linked to the appraisal of safety-reversal, but not threat-reversal, showed a group difference in a region separating the right nucleus accumbens and the putamen.

**Conclusions:** This study advances knowledge of striatal connectivity profiles in OCD, supporting a core role of distinct changes in striatal topology in the expression of symptoms. Collectively, these results encourage studies assessing neural mechanisms driving the dynamic reorganisation of striatal topology and the development of therapies leveraging striato-cortical plasticity.

## Introduction

Obsessive-compulsive disorder (OCD) is a severe mental illness affecting 2-3% of the world population [1]. There is no cure for OCD, and the disorder’s debilitating symptoms are often chronic and can be lifelong [1]. Obsessions and compulsions are core defining symptoms of the disorder, with OCD often presenting alongside co-morbid anxiety disorders, substance abuse and impulse control disorders [2]. While the neural basis of OCD is not fully understood, genetic [3], preclinical [4], neuroimaging [5], and interventional neurosurgical [6, 7] studies have suggested a core deregulation in the activity of the associative-limbic part of the cortico-striatal system [1, 8].

The striatum is a subcortical structure comprising the nucleus accumbens (NAcc), putamen, and caudate. This anatomically and functionally heterogeneous structure is a core entry ganglion of the cortico-basal ganglia-thalamo-cortical loops, and it plays a key role in dynamically integrating and redistributing neural signals across specialised brain regions to generate and orient adaptive behaviours [9]. Converging evidence from multimodal studies in animal models and people with OCD has highlighted altered activity in defined brain circuits encompassing the striatum and the cerebral cortex [8]. Work using animal models presenting OCD-related phenotypes, like excessive grooming, has shown abnormally high activity of medium-sized spiny neurons in a brain region equivalent to the human caudate [10]. This dysregulated activity has been linked to changes in local inhibitory dynamics and altered frontal modulation of striatal microcircuitry activity [11, 12]. At the macroscale, several resting-state and task-based neuroimaging studies have highlighted context-specific changes in the activity and connectivity of frontal and striatal brain network regions [8]. Reduced activity in the cognitive (dorsal-posterior) frontostriatal circuit and increased activity of an affective (ventral-anterior) circuit [1, 13] have been observed in OCD when individuals are at rest [5] or while performing tasks including fear avoidance paradigms [14].

Results from the studies above highlight the likely dynamic nature of the changes in how regions of the striatum are functionally interconnected with cortical territories in OCD compared to control subjects. Changes in the functional topological organisation of the striatum across time may link to fluctuations in the severity of OCD symptoms, identifying new locations for treatment targets (e.g., for neuromodulation) and outcome markers. The functional organisation of the striatum can be measured using the novel concept of functional gradients. Gradients depart from the common delineation of discrete regions and continuously represent topographic changes in connectivity along overlapping organisational axes [15–17]. This approach has uncovered a complementary representation of subcortical regions based on brain functional connectivity, including the striatum [18]. These findings are also in line with results from methods defining dorsal-ventral striatal gradients, aligning them with biological [19, 20] gradients and behaviour [21].

Specifically, mapping functional connectivity gradients allows the characterisation of whole-brain connectivity patterns across striatal voxels. Contrary to standard region-based analyses, this approach permits the definition of continuous variations in the patterns and magnitude of whole-brain connectivity across the striatum. The striatal gradient explaining the most variance in the data is known to capture functional differences between established striatal structures (caudate, NAcc, and putamen) [18]. Thus, our analysis could detect nuanced changes in the functional profile of these structures and how they functionally interconnect with the rest of the OCD brain. A high gradient magnitude in striatal voxels can be used to define locations of rapid changes in how those voxels are functionally coupled with the rest of the brain. Conversely, smaller gradient magnitudes define clusters of relatively homogenous functional connectivity. Changes in striatal magnitude can thus point to defined differences in OCD striatal connectivity profiles that may map onto changes in function. Using this approach, we assessed possible changes in striatal gradients across the resting state and the appraisal of threat or safety stimuli between people with OCD and comparable controls. Assessing task-based changes in striatal gradients allowed testing for possible group differences beyond resting brain activity. Based on previous literature [14], we expected group differences while people engage in the reappraisal of threatening stimuli as safe, but not necessarily in the inverse condition. Differences in gradient magnitudes between these two groups indicate a change in the functional organisation of connectivity between the striatum and the brain. To assess the possible link between changes in striatal functional topology and treatment targets that are known to be effective in OCD, we examined the proximity of OCD-related striatal gradient topology to grey matter clusters [7] and white matter tracts [22] associated with response to deep brain stimulation (DBS) therapy. Further, by taking advantage of this cohort’s longitudinal sampling, we tested whether changes in how resting-state functional gradients differ between OCD and controls related to variations in symptom severity across time.

## Material and Methods

### Participants

The study included 52 people with OCD aged 18-50 (ACTRN12616001687482, see SMaterial). This group was matched with 45 controls [23] (Table 1). Of those 52 OCD participants, 47 completed a follow-up assessment (SMaterial).

**Table 1:** Sample characteristics. Age is reported in years. Characteristics include: the Wechsler Abbreviated Scale of Intelligence (WASI), Yale-Brown Obsessive-Compulsive Scale (Y-BOCS), Hospital Anxiety and Depression scale (HADS), Hamilton Anxiety Rating Scale (HAM-A), Montgomery-Åsberg Depression Rating Scale (MADRS), Obsessional Beliefs Questionnaire (OBQ), and Obsessional Compulsive Inventory Revised (OCIR). Two sample t-tests are used to compare continuous measures, chi-square tests for comparing gender, and Fisher’s exact test for handedness due to small frequencies. ^change is the post-treatment timepoint minus the baseline (divided by baseline Y-BOCS in the case of % change), meaning a negative change indicates clinical improvement.

|  | Controls (N=45) | OCD (N=52) | \|t\|/χ | p |
| --- | --- | --- | --- | --- |
| Age, mean (SD) | 32.5 (8.7) | 30.2 (7.8) | 1.39 | 0.17 |
| Gender, n (%) female | 27 (60.0%) | 29 (55.8%) | 0.05 | 0.83 |
| Handedness, n (%) right | 43 (95.6%) | 44 (84.6%) | - | 0.10 |
| IQ (WASI), mean (SD) | 112.7 (11.3) | 105.9 (12.5) | 2.79 | 0.006 |
| Y-BOCS, mean (SD) | 1.8 (3.0) | 25.4 (5.1) | 27.11 | <0.001 |
| Obsessions | 1.2 (2.3) | 13.8 (4.2) | 17.77 | <0.001 |
| Compulsions | 0.6 (1.6) | 13.3 (2.6) | 28.34 | <0.001 |
| Change in Y-BOCS (N=47)<sup>^</sup> |  | -4.7 (5.2) |  |  |
| % Change in Y-BOCS (N=47)<sup>^</sup> |  | -18.6% (21.2%) |  |  |
| HADS Anxiety, mean (SD) | 4.7 (3.7) | 13.2 (4.6) | 9.86 | <0.001 |
| HADS Depression, mean (SD) | 2.1 (2.3) | 8.2 (4.7) | 7.83 | <0.001 |
| HAM-A, mean (SD) | 2.9 (3.1) | 20.1 (8.8) | 12.45 | <0.001 |
| MADRS, mean (SD) | 3.0 (3.5) | 19.7 (10.6) | 10.11 | <0.001 |
| OBQ, mean (SD) | 116.6 (47.1) | 197.2 (51.4) | 8.01 | <0.001 |
| OCIR, mean (SD) | 5.6 (7.1) | 33.1 (14.6) | 11.54 | <0.001 |

### Neuroimaging data acquisition and analyses

Neuroimaging data were acquired on a 3T scanner (SMaterial). Images were preprocessed using fMRIprep (version 23.2.0) [24] and Nilearn [25] (SMaterial).

### Striatal masks

We used the Melbourne Subcortex Atlas (S1) to isolate the striatum [18], which included the NAcc, putamen, and caudate. We performed functional connectivity (FC) gradient analysis (see below and SMaterial) for the striatum in each hemisphere.

### Striatal functional gradients

Functional gradients describe patterns of changes in the connectivity between the striatum and the rest of the brain. These gradients can be used to map locations having distinct changes in connectivity profiles [18] (details in SMaterial). In brief, a functional connectivity ‘fingerprint’ to the dimensionally reduced whole brain grey matter activity was calculated at each voxel in the striatum (SMaterial, SFigure 1A). For the fear reversal task, we employed generalised psychophysiological interaction (PPI) analysis to estimate task-based connectivity [26, 27], resulting in matrices of coefficient t-statistics (SMaterial).

**Figure 1.**
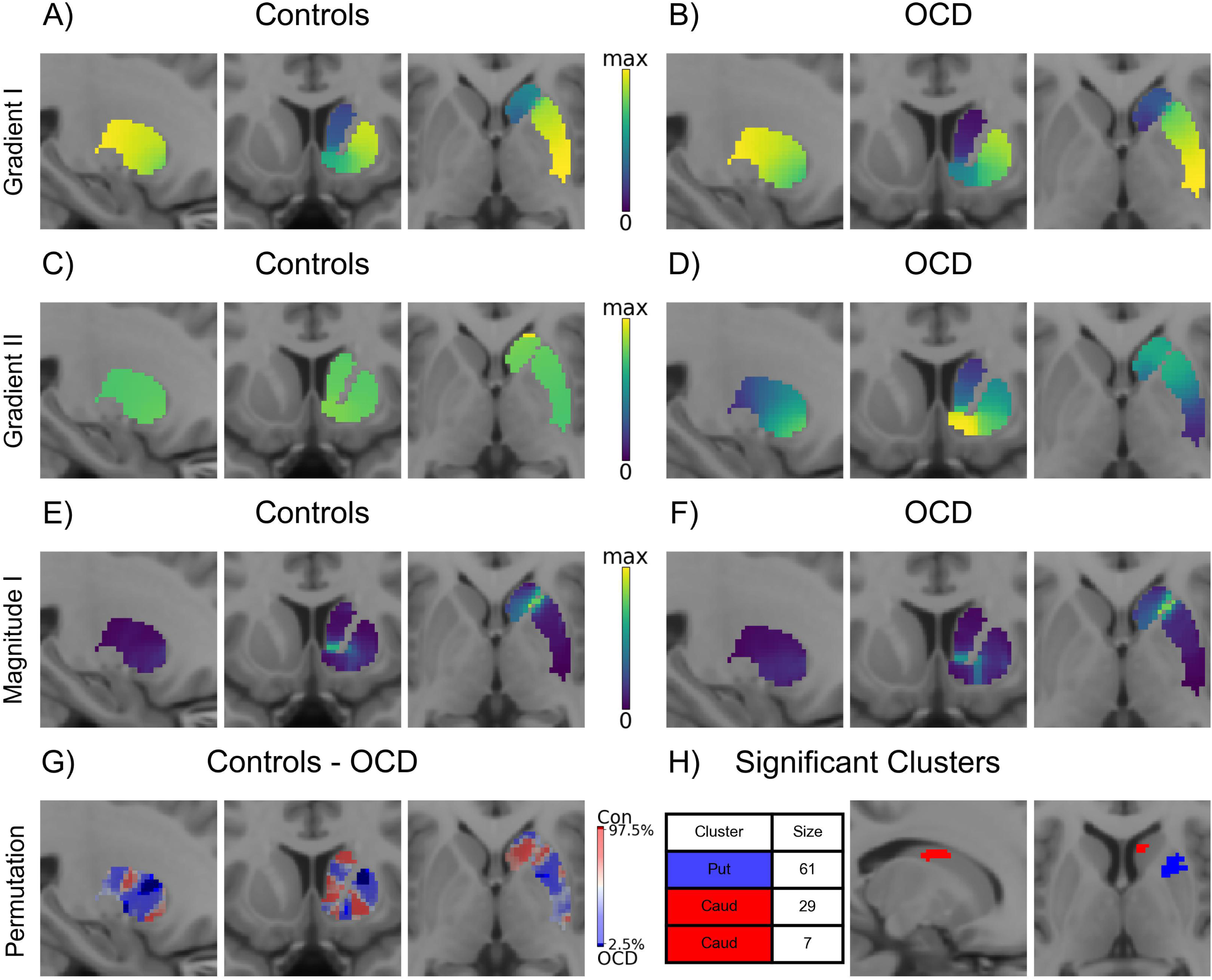
Resting-state right striatal gradients in controls and OCD. A) The panel depicts the topology of the average Gradient I eigenmap for the control group. The variance explained was 20.7%. B) The average Gradient I eigenmap for OCD, explaining 22.9% of the variance. C) The average Gradient II eigenmap for controls explained 6.0% of the variance, while in OCD, the same gradient explained 4.6%. (D). The maximum magnitude for the average Gradient I in controls was 0.806 (E), while it was 0.766 for OCD (F). G) The panel shows percentiles of the differences in magnitude ascribed using group-permutations (uncorrected, Methods). Dark red indicates voxels where the magnitude was higher in Control gradient I while dark blue indicates voxels where the magnitude was higher in OCD gradient I. Imaging slice coordinates for A-G: x = 25, y = 10, z = 0 (MNI space). H) Clusters of voxels with significantly different magnitudes. One large cluster is located in the putamen (Put) and two in the caudate (Caud). Imaging slices in H: x=16, z = 6 (MNI).

The similarity between each striatal voxel in FC fingerprint (rest) or PPI t-statistic (task) were calculated (SMaterial). Connectivity gradients (also termed eigenmaps) and spatial rates of change (gradient magnitudes) were calculated following the approach by Tian, Margulies [18] and Borne, Tian [28]. A Gradient represents a continuous mode of spatial variation in striatal functional connectivity to the whole brain (similar Gradient values indicate similar connectivity profiles to the brain). In contrast, the magnitude represents the degree to which the spatial variation in functional connectivity changes. A higher magnitude at a voxel indicates the striatal FC to the whole brain is changing rapidly across the surrounding voxels (as would occur between distinct substructures), and a lower magnitude at a voxel indicates the FC is very similar to the surrounding voxels.

This method yields N eigenvectors that consecutively explain decreasing variance in the data. The second eigenvector is called Gradient I, while the third is Gradient II (and so on). In line with previous work [17, 24], we considered Gradient I for each of our analyses as this gradient captures the core functional topology of the striatum. We also present Gradient II for rest, as it explains ∼5% of the variance.

Our neuroimaging protocol was optimised for clinical populations, raising questions about the possibility of mapping striatal gradients. Thus, we compared our controls to the average similarity matrix generated using HCP data (n = 812) [18]. The HCP Gradient I was highly comparable to that observed in our control group (SFigure 2; Gradient I spatial correlation of r>0.986), though it was not similar for Gradient II.

**Figure 2.**
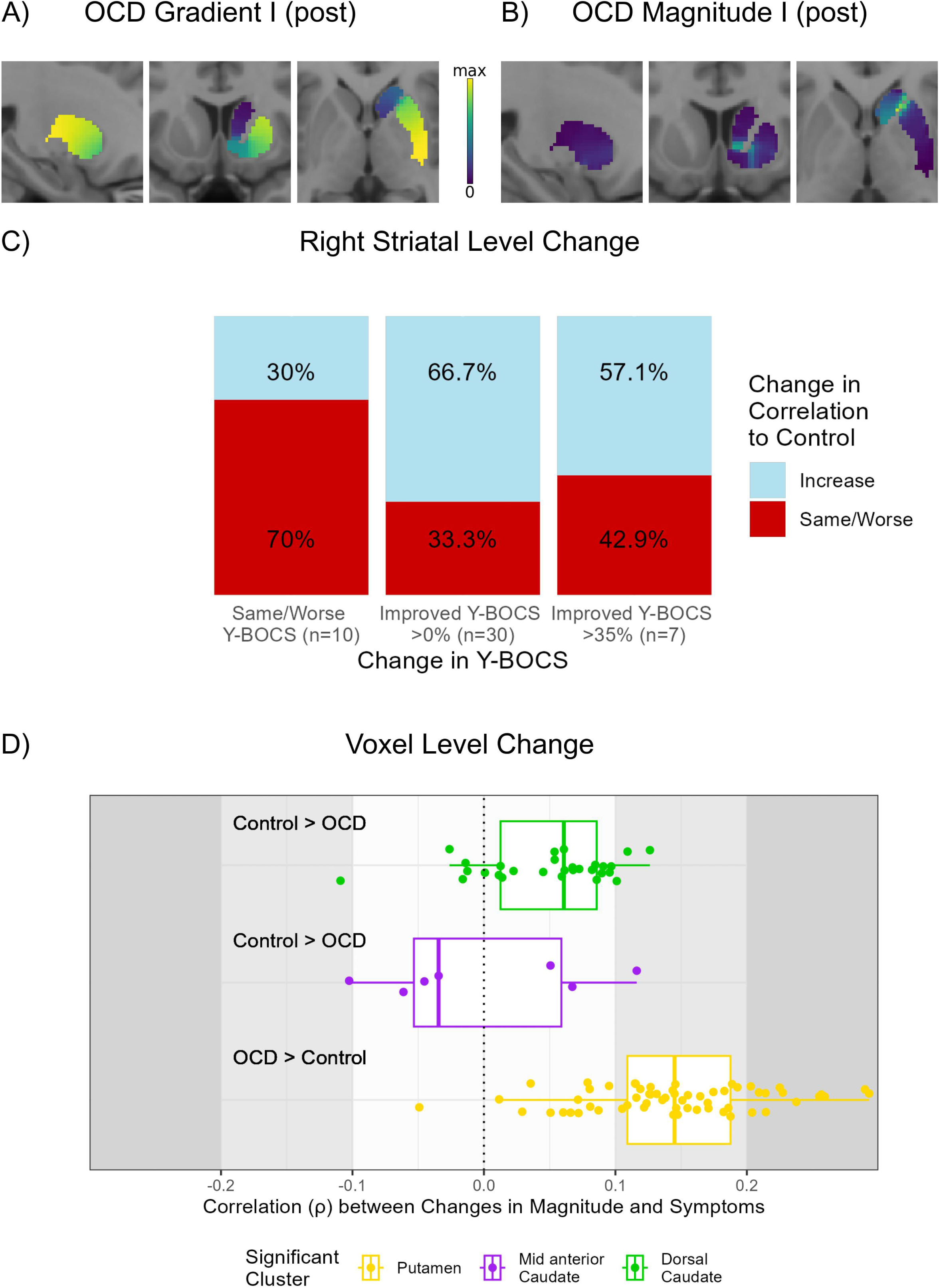
Association between the normalisation of resting state eigenmaps in OCD and changes in symptoms. The average OCD Gradient I at the second time-point explained 25.1% of the variance (A) and showed a maximum magnitude of 0.767 (B). Imaging slice coordinates for (A) and (B): x = 25, y = 10, z = 0 (MNI coordinates). C) Change in the correlation of the OCD individual eigenmaps (Gradient I topology) to the control group average Gradient I by the change in Y-BOCS scores. In the groups with clinical improvement, a higher proportion of individuals Gradient I became more correlated with the Control Gradient I. D) Association between change in magnitude and clinical change in brain voxels belonging to clusters showing a statistically significant group difference (Figure 1H). The association is quantified using the spearman correlation between the change in Y-BOCS (percent change) and the change in magnitude. Voxels in the caudate clusters have a weak association (absolute correlation less than 0.1). On the other hand, voxels in the putamen cluster showed a higher association between the longitudinal normalisation of OCD Gradient 1 and symptom reduction.

### Group differences at rest

For group comparison (SFigure 1C), the group-specific similarity matrices were averaged before the sparse matrix thresholding and the Laplacian were applied. To assess group differences, gradient magnitudes were compared voxel-wise in the striatum through permutation testing (shuffling participants across groups, n=1000, SMaterial). A voxel significant difference was ascribed if the observed percentile was in the top or bottom 2.5% of the null distributions [28]. The voxel-level statistic was complemented by a cluster-extent approach (cluster sizes p<0.05 [29]). Clusters were defined as contiguous voxels in the top or bottom 2.5% of the voxel-wise null distributions (SMaterial).

### Group differences in the threat-safety reversal task

Fear conditioning is thought to be impacted in OCD, whereby neutral stimuli can become conditioned stimuli (CS+ or CS-) when paired with conditioned stimuli (such as internal obsessions) [30]. The threat-safety reversal task is widely used to explore fear-conditioning, with our previous work highlighting cortical and striatal activity linked to flexibly assessing threat levels of external stimuli [23]. Studies have highlighted changes in the striatum activity when people with OCD need to update the threatening or safety nature of stimuli, reflecting a lack of flexible tracking of updated CS+ and CS-contingencies [14]. Group comparison of task conditions was performed similarly to the resting-state comparison, except we used the PPI coefficient t statistics [28] relating to the threat and safety conditions instead of correlation (SFigure 1E). This approach isolates task-evoked changes in striatal gradients over and above rest connectivity [31]. Group differences in gradient magnitudes were assessed, and the same permutation approaches adopted for the rest data were used for the task conditions (SFigure 1F and G, respectively).

### Association between striatal gradient at rest and longitudinal symptom changes

Finally, we assessed if the striatal functional topology of individuals with OCD became more similar to the control topology (average eigenmap) as a function of symptom improvement over time. Active neuromodulation treatment did not differ in outcome from sham intervention in the RCT [32]. Both active and sham interventions resulted in a group-level reduction of symptoms (Table 1).

To assess changes in an individual striatal gradient across time (SFigure 1D), we applied the graph Laplacian to individual similarity matrices at both time points and calculated spatial correlation to the control eigenmap (SMaterial). To assess if a normalisation of OCD individual rest eigenmaps linked to change in Y-BOCS, we compared the number of OCD subjects showing greater eigenmap similarity with controls at follow-up relative to baseline. We tested for a difference in the association with control eigenmaps in three OCD groups: (i) OCD subjects who at follow up showed no improvement or increased symptoms compared to baseline; (ii) subjects who showed small symptoms’ improvement (Y-BOCS changes >0% but <35%), and (iii) subjects who experienced a large improvement in symptoms (Y-BOCS changes ≥35%). The criteria to differentiate group (ii) from group (iii) was in line with [33]. A Fisher’s exact test was used to compare groups.

Some individual eigenmaps had poor correlation and showed markedly different topologies to the group average. Other work has assessed the similarity of individual-level Gradients to the group-level [16]. A previous work has assessed the variability of individual subcortical gradients, noting that individual variation was evident in the striatum [18]. We performed a confirmatory (sensitivity) analysis to assess if the eigenmap normalisation linked to a reduction in OCD symptoms replicated once noisy individual maps were removed via exclusion criteria (SFigure 3).

**Figure 3.**
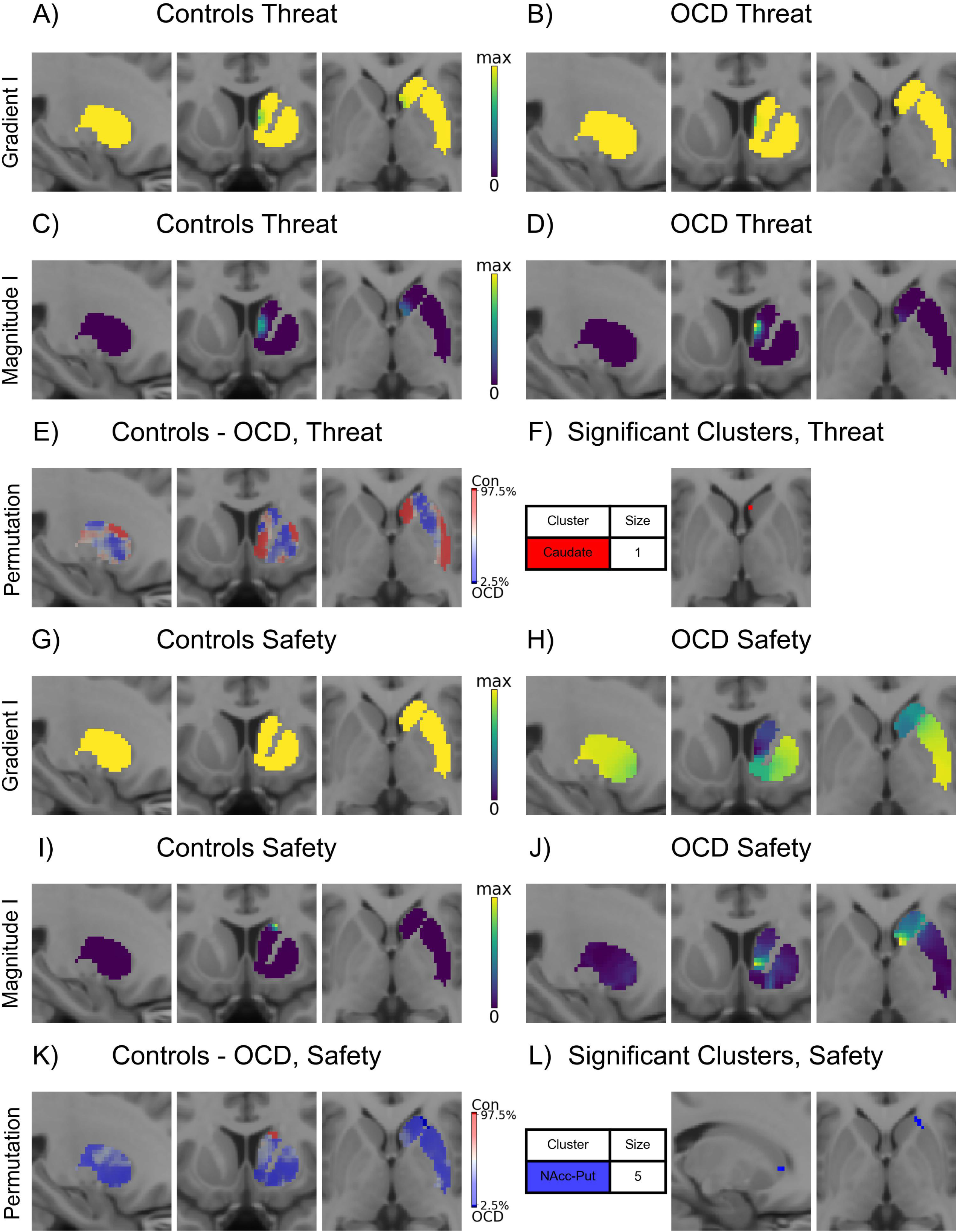
Changes in task-specific functional gradients in controls and OCD. The figure presents the results for the right striatum. A) The control group Gradient I eigenmap for the threat-reversal condition. This gradient explained 18.0% of the variance. B) The panel shows the OCD Gradient I eigenmap for the threat-reversal condition. In OCD, this gradient explained 24.0% of the variance. C) Average gradient I magnitude for the threat-reversal condition in controls. The maximum magnitude was 3.948. D) Average gradient I magnitude of the threat-reversal condition in OCD. The maximum magnitude was 3.979. E) Percentiles of the voxel-wise group magnitude difference during threat-reversal condition (high threshold p<0.05, uncorrected). Dark red indicates significant voxels with higher magnitude values in the Control compared to OCD. Conversely, dark blue voxels showed higher magnitude in OCD Gradient I compared to the control Gradient I. Imaging slice coordinates for A-E: x = 25, y = 10, z = 0 (MNI space) F) Cluster of a single voxel with significantly different magnitude in the caudate. Imaging slice in F: z=2 (MNI). G) Representation of the control group average Gradient I eigenmap associated to the safety-reversal task condition. The variance explained was 10.5%. H) The average Gradient I eigenmap for the safety condition for OCD, which explained 10.5% of the variance. I) The average Gradient I magnitude for the safety-reversal condition in controls. The maximum magnitude was 0.734. J) The safety average Gradient I magnitude in OCD, showing a maximum magnitude of 0.982. K) Percentiles of the voxel-wise group magnitude difference linked to the safety-reversal condition isolated using a voxel-level high threshold of p<0.05 (uncorrected). Dark red indicates voxels in which the magnitude was higher in the control Gradient I. Conversely, dark blue indicates that the magnitude was higher in OCD Gradient I. Imaging slice coordinates for G-K: x = 25, y = 10, z = 0 (MNI space) L) Cluster of voxels with significantly different magnitudes between the NAcc and putamen (Put). Imaging slice in L: x = 18, z = 0 (MNI).

We also assessed changes in gradient magnitude in voxels found to be significantly different between the groups. Voxel-level association was assessed with Spearman correlation (change in magnitude at significant voxels and change in Y-BOCS).

## Results

### Group differences in gradients at rest

In both groups, Gradient I captured the distinct connectivity profiles of the caudate, NAcc, and putamen with the rest of the brain, (Figure 1A-B). These results are in line with what was observed by [18] (SFigure 2). On the other hand, Gradient II differentiated the NAcc from the other structures, though less evidently in the controls compared to the participants with OCD (Figure 1C-D). The variance explained by Gradient I was 20.7% and 22.9% for control and OCD, respectively. Gradient II only explained 4.6% of the variance in OCD and 6.0% in controls. Thus, the following analyses focus on Gradient I.

The peaks of the Gradient I magnitudes in both groups occur on either side of the NAcc, separating this structure from the caudate and putamen (Figure 1E-F). A similar pattern of results was observed in the left striatum (SFigure 4). Voxel permutation testing highlighted several regions in the putamen and caudate with higher and lower gradient magnitudes in OCD relative to controls (Figure 1G and SFigure 5). Follow-up cluster-extent analysis defined clusters showing a statistically significant difference in magnitude between groups (p<0.05, Figure 1H). A large cluster overlapping the ventro-anterior portion of the putamen showed a higher Gradient I magnitude in OCD (blue colour in Figure 1H). Conversely, two smaller clusters in the caudate (dorsal peak and anterior-ventral; red colour in Figure 1H and SFigure 5) showed lower Gradient I magnitude in OCD than controls. These results suggest differences in how the OCD dorsal striatum interacts with the rest of the brain.

Next, we assessed if clusters showing a group effect were located near known targets for deep brain stimulation for OCD [7, 22]. Results showed no overlap or adjacent spatial relationships (SFigure 6).

### Association between clinical and resting-state changes across time

The characteristic caudate to putamen pattern of Gradient I was also observed in the OCD group at follow-up (second time point; Figure 2A). As observed at baseline, voxels with peak magnitude values delineated traditional boundaries between the NAcc, the caudate, and the putamen (Figure 2B). We investigated a possible association between baseline OCD symptom severity and OCD deviations from the control striatal functional topology. Results showed a very weak association (r<0.03) at the striatum level, and week associations at the voxel level (SFigure 7).

Between the two time points, many individuals with OCD showed clinical improvement, reflected in a reduction in their total Y-BOCS scores. We hypothesized that the decrease in total Y-BOCS scores linked to changes in Gradient I topology. In particular, we predicted that the closer the OCD topology changed towards the topology of controls, the stronger the clinical improvement. This relationship was difficult to assess along a continuum due to the noisiness of individual Gradients (SFigures 8 and 9). Thus, we categorised the symptom improvement and binarized the change in correlation to control topology into a simple increase (OCD Gradient 1 increased its similarity to the control Gradient 1) or decrease (OCD Gradient 1 decreased its similarity to the control Gradient 1). In line with our prediction, we observed a numerical reduction in symptoms as a function of improved similarity between OCD individuals’ Gradient I and the control gradient. However, this effect did not reach statistical significance (Figure 2C, p = 0.12, Fisher’s exact test; Figure 2C).

Several individuals’ Gradient I varied radically from the group average (SFigure 8 for examples). Thus, we performed a sensitivity analysis excluding individuals with OCD with a markedly different Gradient I topology. Results from this sensitivity analysis were not significant (p = 0.22, Fisher’s exact test) but they supported the hypothesis that participants with improved Y-BOCS showed a higher proportion of normalization of Gradient I topology (SFigure 3). Considering the change in Y-BOCS scores and the change in correlation to control Gradient I along a continuum revealed only a weak correlation (SFigure 9).

We also explored whether changes in the Gradient I magnitude in the clusters identified as different between groups (Figure 1H) were associated with fluctuations in symptom severity (SFigure 10). While magnitude changes in the caudate clusters had very weak associations with changes in OCD symptoms (|r|<0.1; Figure 2D), voxels in the putamen cluster (which had greater magnitude in OCD vs. Controls) showed slightly stronger positive associations (i.e., greater normalization of magnitude associated with greater symptom improvements).

### Task conditions

Next, we examined whether group differences in task-related striatal gradients occurred in OCD. Task-related gradients represent change above background patterns of connectivity and may only comprise circumscribed striatum regions. We present task-evoked changes in the first two gradients (SFigure 11), but only assessed group differences in the first gradient as the variance explained by Gradient II was small (below).

The threat-reversal Gradient I for controls explained 18.0% of the variance (Figure 3A), while it explained 24.0% in OCD (Figure 3B). Gradient II explained 8.2% and 6.7% of the variance for controls and OCD, respectively (SFigure 11). The bulk of threat-reversal-induced changes in striatal topology was circumscribed (Figure 3A and B). Accordingly, in both groups, the Gradient I magnitude peaked in the mid-caudate (Figure 3C and D), with magnitude close to zero elsewhere. Permutation tests of Gradient I magnitude (Figure 3E) showed group differences in only one voxel (Figure 3F) in the right caudate.

In the safety reversal condition, Gradient I explained 10.5% of the variance in both the control (Figure 3G) and the OCD group (Figure 3H). Gradient II (SFigure 11) explained variance was 2.5% for controls and 3.8% for OCD. In controls, Gradient I (Figure 3G) separated the dorsal tip of the caudate from the rest of the striatum (Figure 3I), though the spatial variation was almost none-existent across most of the striatum (variation in only a few voxels on the face of the dorsal caudate). However, in OCD, this gradient showed a caudate to NAcc to putamen topology (Figure 3H), with large gradient magnitudes on either side of the NAcc (Figure 3J). Voxel-wise group differences suggested a difference in Gradient I magnitude throughout the NAcc and putamen (dark blue in Figure 3K). Accordingly, a single cluster crossing the NAcc and putamen had a significantly higher Gradient I magnitude in OCD than controls (Figure 3L). This difference, emerging on top of a stronger functional segregation of striatal structures in OCD relative to controls, suggests a different engagement of the striatum during the appraisal of safety.

While the Safety-Threat Reversal task was also performed longitudinally, the individual gradients calculated using PPI coefficient t statistics were too noisy for a within-subject comparison.

## Discussion

The striatum is a key subcortical hub that receives information from and sends signals to other brain structures [34–36]. Recent studies have shown that functional organisations can be described compactly via gradients [15, 18]. The ability to describe the organisation of the striatum using continuous gradients allows a holistic appraisal and systems-level explanation of how functional interactions between the striatum and the whole brain link to OCD [37]. These interactions map onto a striatal topological gradient defined by functionally integrated brain regions [38]. Evidence from preclinical [4, 39], human neuroimaging [5, 8, 40, 41], and neurosurgery [22, 42] studies highlight changes in the functional interplay between striatal regions and other brain regions, supporting variations in the functional topology of the OCD striatum. Here, we used in vivo mapping of functional gradients of the human brain [18] to assess changes in the striatal resting-state functional topology in individuals with clinical OCD. This novel method captures the functional organisation of whole brain connectivity through the striatum, expanding on previous region of interest approaches. Further, we tested whether predicted variations in the resting topology of the striatum generalise to behavioural contexts relevant for OCD (flexible processing of threatening and safe stimuli, [14, 43]). Finally, we assessed whether longitudinal changes in striatal functional gradients may be related to fluctuations in OCD symptoms. Our results suggest context-specific changes in the functional topology of the striatum that enable OCD and may carry clinical value. This new knowledge on the atypical macroscale organisation of interactions between the striatum and the rest of the brain motivates future work assessing the biological underpinnings of these deregulations [44], the link between structure-function-behaviour [45, 46], the use of these advances for biomarker development, and the possible normalisation of these couplings via therapeutic interventions.

At rest, the topology and explained variance of the two major striatal gradients were similar between OCD and controls. However, OCD showed distinct clusters of significant changes in gradient magnitude in the putamen and caudate. These findings aligned with several preclinical and clinical studies suggesting changes in distinct frontostriatal circuits [47] in the resting state [1, 13, 48]. The putamen cluster shows increased heterogeneity in the connectivity with the rest of the brain. These results align with previous analyses of frontostriatal circuits activity at rest, showing core deregulation in connectivity between the putamen and the lateral prefrontal cortex in OCD [40]. Meanwhile, the caudate clusters showing decreased gradient magnitude in OCD highlight a decreased heterogeneity in how these regions interact with the brain. In line with the behavioural phenotype of OCD, these clusters are part of the dorsal striato-cortical system supporting appropriate planning [49] and goal-directed behaviour [50]. The clinical significance of resting-state changes in striatal Gradient I is supported by the suggestion that adjustments in individual striatal topology towards the group-average topology of controls link to an improvement in OCD symptoms (although not significantly, assuming a type I error of 5%). These findings put forward the hypothesis that OCD is underpinned by reversible changes in striatal connectivity, encouraging the development of targeted therapies leveraging resting state striato-cortical plasticity. Functional connectivity gradients are proposed as a novel biomarker approach, offering better phenotypic prediction than traditional parcellation-based methods [17, 51].

The appraisal of threat and safety was related to task-evoked changes in the first functional gradient in OCD relative to controls. Generally, our results align with the observation that task-related functional architecture is mainly shaped by resting state connectivity, with limited connectivity changes evoked by a given task [52]. While minimal group differences were observed for the threat condition, analyses revealed a cluster of significant changes in the gradient magnitude between the right putamen and the NAcc when individuals appraised safety, indicating greater functional segregation between the two structures in OCD. However, contrary to what was expected based on previous results adopting a threat-safety appraisal paradigm [14], this difference was not observed in the left hemisphere. Our previous analysis of this dataset found no group differences between patterns of activity linked to threat and safety reversal within core regions engaged by these mental processes [23]. By adopting an analysis that captures continuous topographic changes in connectivity, the current results add to previous knowledge by supporting the role of the striatum, in addition to the ventromedial prefrontal cortex, in the altered appraisal of safety in OCD [14]. These advances progress our understanding of the neural basis of altered behaviour in OCD, encouraging the development of context-specific therapies to restore the neural substrate supporting the adaptive assignment of safety to external stimuli. Future work could explore functional gradients in other task contexts related to OCD, such as a task probing the ability to switch from habitual to goal-directed behaviour [53].

Some limitations need to be considered when interpreting the results of this study. First, the mapping of functional gradients in the striatum using fMRI is vexed by a low signal-to-noise [18]. Moreover, the duration of the rest acquisition and the number of task events were limited to enhance feasibility and reduce attrition. These limitations resulted in data losses and may have precluded the detection of more subtle changes in the functional topology of the striatum in OCD. Future studies could test the feasibility of performing precision functional mapping in people with OCD [54], facilitating the reliable detection of OCD-induced changes in striatal topology and the establishment of biomarkers that could be used to personalise interventions and assess treatment efficacy in clinical trials. We note that the IQ in healthy controls was above-average, relative to the average IQ in the OCD group. Thus, replication studies are required to confirm the observed results in an IQ-matched sample. Follow-up studies with larger sample sizes should also explore potential changes in the striatal functional topology in more diverse clinical groups, including younger participants who may be more treatment-naïve and individuals with comorbid conditions.

The current study highlights distinct and context-specific changes in the functional topology of the striatum in individuals diagnosed with OCD. These results align with the suggested key role of altered striato-cortical activity in OCD and with evidence from preclinical and clinical studies indicating that the normalisation of local striatal dynamics and its outputs relates to clinical improvements. The observed plasticity of the striatum functional topology indicate that OCD is related to functional deregulations that could, at least in part, be corrected with targeted interventions.

## Data Availability

All data produced in the present study are available upon reasonable request to the authors

## References

1. Stein, D.J., et al., Obsessive–compulsive disorder. Nature Reviews Disease Primers, 2019. 5(1): p. 52.

2. Ruscio, A.M., et al., The epidemiology of obsessive-compulsive disorder in the National Comorbidity Survey Replication. Molecular Psychiatry, 2010. 15(1): p. 53–63.

3. Stewart, S.E., et al., Genome-wide association study of obsessive-compulsive disorder. Mol Psychiatry, 2013. 18(7): p. 788–98.

4. Ahmari, S.E., et al., Repeated Cortico-Striatal Stimulation Generates Persistent OCD-Like Behavior. Science, 2013. 340(6137): p. 1234–1239.

5. Naze, S., et al., Mechanisms of imbalanced frontostriatal functional connectivity in obsessive-compulsive disorder. Brain, 2022. 146(4): p. 1322–1327.

6. Figee, M., et al., Deep brain stimulation restores frontostriatal network activity in obsessive-compulsive disorder. Nat Neurosci, 2013. 16(4): p. 386–7.

7. Meyer, G.M., et al., Deep Brain Stimulation for Obsessive-Compulsive Disorder: Optimal Stimulation Sites. Biological Psychiatry, 2024. 96(2): p. 101–113.

8. Robbins, T.W., M.M. Vaghi, and P. Banca, Obsessive-Compulsive Disorder: Puzzles and Prospects. Neuron, 2019. 102(1): p. 27–47.

9. Alexander, G.E. and M.D. Crutcher, Functional architecture of basal ganglia circuits: neural substrates of parallel processing. Trends Neurosci, 1990. 13(7): p. 266–71.

10. Piantadosi, S.C., et al., Hyperactivity of indirect pathway-projecting spiny projection neurons promotes compulsive behavior. Nature Communications, 2024. 15(1): p. 4434.

11. Burguière, E., et al., Striatal circuits, habits, and implications for obsessive–compulsive disorder. Current Opinion in Neurobiology, 2015. 30: p. 59–65.

12. Burguière, E., et al., Optogenetic stimulation of lateral orbitofronto-striatal pathway suppresses compulsive behaviors. Science, 2013. 340(6137): p. 1243–6.

13. Shephard, E., et al., Toward a neurocircuit-based taxonomy to guide treatment of obsessive-compulsive disorder. Mol Psychiatry, 2021. 26(9): p. 4583–4604.

14. Apergis-Schoute, A.M., et al., Neural basis of impaired safety signaling in Obsessive Compulsive Disorder. Proceedings of the National Academy of Sciences, 2017. 114(12): p. 3216–3221.

15. Margulies, D.S., et al., Situating the default-mode network along a principal gradient of macroscale cortical organization. Proceedings of the National Academy of Sciences, 2016. 113(44): p. 12574–12579.

16. Haak, K.V., A.F. Marquand, and C.F. Beckmann, Connectopic mapping with resting-state fMRI. NeuroImage, 2018. 170: p. 83–94.

17. Oldehinkel, M., et al., Gradients of striatal function in antipsychotic-free first-episode psychosis and schizotypy. Transl Psychiatry, 2023. 13(1): p. 128.

18. Tian, Y., et al., Topographic organization of the human subcortex unveiled with functional connectivity gradients. Nature Neuroscience, 2020. 23(11): p. 1421–1432.

19. Haber, S.N., J.L. Fudge, and N.R. McFarland, Striatonigrostriatal pathways in primates form an ascending spiral from the shell to the dorsolateral striatum. J Neurosci, 2000. 20(6): p. 2369–82.

20. Oldehinkel, M., et al., Mapping dopaminergic projections in the human brain with resting-state fMRI. Elife, 2022. 11.

21. Marquand, A.F., K.V. Haak, and C.F. Beckmann, Functional corticostriatal connection topographies predict goal directed behaviour in humans. Nat Hum Behav, 2017. 1(8): p. 0146.

22. Li, N., et al., A unified connectomic target for deep brain stimulation in obsessive-compulsive disorder. Nature Communications, 2020. 11(1): p. 3364.

23. Hearne, L.J., et al., Revisiting deficits in threat and safety appraisal in obsessive-compulsive disorder. Human Brain Mapping, 2023. 44(18): p. 6418–6428.

24. Esteban, O., et al., fMRIPrep: a robust preprocessing pipeline for functional MRI. Nature Methods, 2019. 16(1): p. 111–116.

25. Abraham, A., et al., Machine learning for neuroimaging with scikit-learn. Frontiers in Neuroinformatics, 2014. 8.

26. Friston, K.J., et al., Psychophysiological and modulatory interactions in neuroimaging. Neuroimage, 1997. 6(3): p. 218–29.

27. McLaren, D.G., et al., A generalized form of context-dependent psychophysiological interactions (gPPI): a comparison to standard approaches. Neuroimage, 2012. 61(4): p. 1277–86.

28. Borne, L., et al., Functional re-organization of hippocampal-cortical gradients during naturalistic memory processes. Neuroimage, 2023. 271: p. 119996.

29. Lindquist, M.A. and A. Mejia, Zen and the art of multiple comparisons. Psychosom Med, 2015. 77(2): p. 114–25.

30. Cooper, S.E. and J.E. Dunsmoor, Fear conditioning and extinction in obsessive-compulsive disorder: A systematic review. Neuroscience & Biobehavioral Reviews, 2021. 129: p. 75–94.

31. O’Reilly, J.X., et al., Tools of the trade: psychophysiological interactions and functional connectivity. Soc Cogn Affect Neurosci, 2012. 7(5): p. 604–9.

32. Cocchi, L., et al., Effects of transcranial magnetic stimulation of the rostromedial prefrontal cortex in obsessive–compulsive disorder: a randomized clinical trial. Nature Mental Health, 2023. 1(8): p. 555–563.

33. Mataix-Cols, D., et al., Operational Definitions of Treatment Response and Remission in Obsessive-Compulsive Disorder Capture Meaningful Improvements in Everyday Life. Psychotherapy and Psychosomatics, 2022. 91(6): p. 424–430.

34. Spiliotis, K., et al., Data-driven and equation-free methods for neurological disorders: analysis and control of the striatum network. Frontiers in Network Physiology, 2024. 4.

35. Straub, C., et al., Principles of Synaptic Organization of GABAergic Interneurons in the Striatum. Neuron, 2016. 92(1): p. 84–92.

36. Aron, A.R. and R.A. Poldrack, Cortical and Subcortical Contributions to Stop Signal Response Inhibition: Role of the Subthalamic Nucleus. The Journal of Neuroscience, 2006. 26(9): p. 2424–2433.

37. Vos de Wael, R., et al., BrainSpace: a toolbox for the analysis of macroscale gradients in neuroimaging and connectomics datasets. Communications Biology, 2020. 3(1): p. 103.

38. Basile, G.A., et al., Striatal topographical organization: Bridging the gap between molecules, connectivity and behavior. Eur J Histochem, 2021. 65(s1).

39. Mondragón-González, S.L., C. Schreiweis, and E. Burguière, Closed-loop recruitment of striatal interneurons prevents compulsive-like grooming behaviors. Nature Neuroscience, 2024. 27(6): p. 1148–1156.

40. Harrison, B.J., et al., Altered Corticostriatal Functional Connectivity in Obsessive-compulsive Disorder. Archives of General Psychiatry, 2009. 66(11): p. 1189–1200.

41. Park, J., et al., Functional Connectivity of the Striatum as a Neural Correlate of Symptom Severity in Patient with Obsessive-Compulsive Disorder. Psychiatry Investig, 2020. 17(2): p. 87–95.

42. Provenza, N.R., et al., Disruption of neural periodicity predicts clinical response after deep brain stimulation for obsessive-compulsive disorder. Nature Medicine, 2024. 30(10): p. 3004–3014.

43. Milad, M.R., et al., Deficits in Conditioned Fear Extinction in Obsessive-Compulsive Disorder and Neurobiological Changes in the Fear Circuit. JAMA Psychiatry, 2013. 70(6): p. 608–618.

44. Müller, E.J., et al., Core and matrix thalamic sub-populations relate to spatio-temporal cortical connectivity gradients. Neuroimage, 2020. 222: p. 117224.

45. Shine, J.M., et al., Human cognition involves the dynamic integration of neural activity and neuromodulatory systems. Nat Neurosci, 2019. 22(2): p. 289–296.

46. Paquola, C., et al., Microstructural and functional gradients are increasingly dissociated in transmodal cortices. PLOS Biology, 2019. 17(5): p. e3000284.

47. Haber, S.N., Corticostriatal circuitry. Dialogues in Clinical Neuroscience, 2016. 18(1): p. 7–21.

48. Hou, J., et al., Morphologic and functional connectivity alterations of corticostriatal and default mode network in treatment-naïve patients with obsessive-compulsive disorder. PLoS One, 2013. 8(12): p. e83931.

49. van den Heuvel, O.A., et al., Frontal-striatal dysfunction during planning in obsessive-compulsive disorder. Arch Gen Psychiatry, 2005. 62(3): p. 301–9.

50. Vaghi, M.M., et al., Specific Frontostriatal Circuits for Impaired Cognitive Flexibility and Goal-Directed Planning in Obsessive-Compulsive Disorder: Evidence From Resting-State Functional Connectivity. Biol Psychiatry, 2017. 81(8): p. 708–717.

51. Hong, S.-J., et al., Toward a connectivity gradient-based framework for reproducible biomarker discovery. NeuroImage, 2020. 223: p. 117322.

52. Cole, Michael W., et al., Intrinsic and Task-Evoked Network Architectures of the Human Brain. Neuron, 2014. 83(1): p. 238–251.

53. Gillan, C.M., et al., Disruption in the balance between goal-directed behavior and habit learning in obsessive-compulsive disorder. Am J Psychiatry, 2011. 168(7): p. 718–26.

54. Gordon, E.M., et al., Precision Functional Mapping of Individual Human Brains. Neuron, 2017. 95(4): p. 791–807.e7.

