## Supplementary Material for "Altered striatal functional gradients in obsessive-compulsive disorder"

**Supplementary Text**

*Sample characteristics*

As part of a registered randomised-controlled clinical trial into the effects of non-invasive brain stimulation on brain activity and symptoms of OCD, people with a clinical diagnosis of OCD and controls were recruited from across Australia. Sample characteristics, including medication use and co-morbidities, have been reported previously [1-3]. Briefly, OCD participants were recruited if they presented with a clinical diagnosis of OCD, as assessed via the Yale Brown Obsessive Compulsive Scale (Y-BOCS [4]), for at least 12 months and no changes in pharmaceutical treatment in the prior month. The diagnosis of OCD was confirmed by a board-certified psychiatrist (B.B or M.B). Inclusion criteria for all participants included age between 18 and 50 years, no history of psychiatric disorders, manic episodes, suicide attempts, traumatic head injuries, neurological disorders, substance or alcohol abuse disorders, alcohol or drug misuse, and no contraindications to MRI. The recruited individuals with OCD typically had mild to moderate anxiety (mean (SD) HAM-A of 19.9 (8.8)) and mild to moderate depression (mean (SD) MADRS of 19.6 (10.4)) [3]. Over half of the individuals were using at least one SSRI (26/50, 52%), 12 were on at least one antipsychotic (24%), and 7 were on at least one benzodiazepine (14%). See Cocchi, Naze [3] for full medication summary.

As reported previously [3], the OCD participants were randomised to receive either continuous theta burst stimulation of the frontal pole via transcranial magnetic stimulation or sham, though no effects of the trial intervention were observed.

Of those 52 OCD participants, 47 completed an assessment and MRI scan at a follow-up timepoint (about 4 weeks following the baseline clinical assessments and MRI session). The change in the primary outcome (Y-BOCS) ranged from a decrease of 18 to an increase of 4.

The local ethics committee approved all procedures (P2253), and all participants signed an informed written consent.

*Neuroimaging data acquisition*

Data were acquired on a 3 Tesla Siemens Prisma MR scanner at the Herston Imaging Research Facility, Brisbane. Whole brain EPI had a voxel size = 2x2x2 mm^3^, TR = 810 ms, and a multiband acceleration factor = 8. Participants underwent a 12-minute eyes-open resting state acquisition followed by a threat-safety reversal task [2]. The fear reversal task lasted 17 min (1227 volumes), and the resting state was 11.9 min (880 volumes). Briefly, in this task, a neutral stimulus (conditioned stimulus [CS]) is differentially conditioned with an aversive auditory event (unconditioned stimulus [US]). The task comprises a habituation, a conditioning, and a reversal phase. During the habituation phase, each neutral stimulus (a blue or yellow sphere shown on a black background for 2 s) was presented five times without the US. During conditioning, the US was presented with one of the coloured spheres (blue or yellow) but not the other. This resulted in CS+ and CS− stimuli. The initial CS+ colour learned in the conditioning phase was counterbalanced across individuals. The US–CS pairing was switched without informing individuals in the reversal phase. The conditioning and reversal phases comprised 5 trials of CS+ paired with the US, 10 trials of the CS+ alone, and 10 trials of the CS−, with the stimuli presented pseudo-randomly to avoid two trial types being successively presented. A white fixation cross lasting 12 s was presented between stimulus trials. We analysed two contrasts from this task associated with flexibly assessing the threat level of stimuli: threat reversal (reversal CS+ > conditioning CS-) and safety reversal (reversal CS- > conditioning CS+). For simplicity, we will hereafter refer to these contrasts as threat and safety, respectively.

Structural brain images were acquired for preprocessing (voxel size = 1x1x1 mm^3^, TR = 1900 ms). Anterior-to-posterior and posterior-to-anterior spin echo fieldmaps were also acquired for preprocessing.

*Analyses*

*Neuroimaging Preprocessing*

The functional brain images were preprocessed using a combination of fMRIprep (version 23.2.0) [5] and Nilearn [6] (Supplementary Material). Images were skull stripped, corrected for susceptibility distortions, coregistered to the anatomical image and slice time corrected. The resulting functional brain images were resampled to the MNI152NLin6Asym 2x2x2 mm^3^ isotropic template space. Both task and resting-state data were denoised using Nilearn with a standard pipeline [7]. Timeseries were demeaned. The following parameters were included in a regression model: 24 head-motion parameters (six motion parameters, their temporal derivatives and quadratics of all regressors), the average white matter and cerebrospinal fluid signals, and cosine transformation basis regressors. This regression model also included the mean global signal for resting-state data. As done in [8] and [9], the images were subsequently spatially smoothed with a 6 mm FWHW Gaussian filter. The Wishart filter [10] was applied in line with Tian, Margulies [8] to further reduce the noise.

*Psychophysiological interactions*

Psycho-physiological interaction (PPI) analysis was performed using a combination of Nilearn [6] and custom python scripts [11]. Timeseries from each subcortical voxel were treated as seeds (T_task_=1227 timepoints). For each seed a separate PPI regression analysis was performed, with the whole brain PCA timeseries were as targets. Each regression model included the seed timeseries, task onsets convolved with the hemodynamic response function and the psycho-physiological interaction term. A T-1 series of interaction term β coefficient t statistics from each regression model corresponding to the PPI term of interest (safety- and fear-reversal) filled a single row of the resulting connectivity matrix in each condition (N rows for each of N subcortical voxels, by T-1 columns for the series of coefficients, Supplementary Figure 12). The GLM coefficients capture the changes in the BOLD signal that can be explained by the task stimuli, above the spontaneous (resting state) fluctuations in the BOLD signal. Given the relatively rapid event design, the PPI term was created after deconvolving the signal using ridge regression [12], implemented as per [11].

*Striatal functional gradients*

Principle components analysis (PCA) was used to reduce each participant's time series (*T* time points) of the *M* grey matter voxels in the grey matter mask (whole brain grey matter mask of 164360 voxels provided by Tian, Margulies [8]) to give a *T* by *T-1* matrix. Then to compute the functional connectivity (FC), we calculated the correlation between the *T* time series in the *N* subcortical voxels and the *T-1* principal components, resulting in an *N* by *T-1* correlation (i.e., FC) matrix (Supplementary Figure 1A).

For the fear reversal task, we employed generalised psychophysiological interaction (PPI) analysis to estimate task-based connectivity [13, 14]. PPI aims to isolate changes in connectivity specifically related to task conditions, above and beyond i) changes in functional connectivity due to spontaneous (intrinsic) fluctuations and ii) co-activations between brain regions. In brief, for each of the *N* subcortical voxels (“seeds”) a regression matrix was constructed that included i) the seed regions timeseries, ii) the task condition event regressor, and iii) the interaction between the two. Deconvolution and mean-centring of the task regressor were employed in line with best practice [14]. The targets in the regression model were the *T-1* principal components representing whole brain timeseries. Thus, the final result was a *condition* by *N* by *T-1* matrix of interaction term $\beta$coefficient *t*-statistics. Conditions were contrasted to generate a task connectivity matrix related to *Threat* and *Safety Reversal* processing (defined above).

Striatal similarity matrices were calculated between sub-cortical voxels (Supplementary Figure 1B), whereby the similarity (i.e. $\eta^{2}$ coefficient) between the *i^th^* and *j^th^* voxels (*i* and *j* in 1 to N) in the correlation matrix (rest) or matrix of *t-*statistics (task conditions) was calculated. This gave an *N*-by-N symmetric similarity matrix.

Connectivity gradients (also termed eigenmaps) and spatial rates of change (gradient magnitudes) were calculated following the approach by Tian, Margulies [8] and Borne, Tian [9]. Before the graph Laplacian was applied, the individual or group-averaged similarity matrices were converted to sparse matrices with a similarity threshold defined as the minimum value to obtain a fully connected graph. The spatial Laplacian gave *N* eigenvectors. The first eigenvector is a constant and not considered for analysis. The second eigenvector is called Gradient I, while the third is Gradient II (and so on). In line with previous work [8, 9], we considered Gradient I for each of our analyses as this gradient captures the core functional topology of the striatum. As it explains about 5% of the variance, we also present Gradient II for rest. See Supplementary Figure 13 for the decrease in variance explained by successive gradients.

Group differences in gradients

For group comparison (Supplementary Figure 1C), the group-specific similarity matrices were averaged before the sparse matrix thresholding and the Laplacian were applied. To assess differences between the groups, gradient magnitudes were compared at each region in the striatum through permutation testing (shuffling participants in each group for n=1000 permutations). After shuffling, the graph thresholding and Laplacian were computed on the permuted group-average similarity matrix. Shuffling the participants between the groups kept the spatial relationship of the eigenmaps consistent, allowing the testing of the null hypothesis that gradient magnitudes at each voxel are equal between groups. A voxel significant difference was ascribed if the observed percentile was in the top or bottom 2.5% of the null distributions for each voxel [9]. Due to the challenge of appropriately accounting for multiple tests, the voxel-level statistic was complemented by a cluster-extent permutation approach assessing the likelihood that the identified voxel cluster sizes were significant at p<0.05 [15]. Clusters were defined as contiguous voxels that were either all in the top 2.5% or all in the bottom 2.5% of the voxel-wise null distributions.

*Association between striatal functional topology at rest and longitudinal changes in symptoms*

To assess changes in an individual striatal gradient across time (Supplementary Figure 1D), we applied the graph Laplacian to individual similarity matrices at both time points. Each resulting eigenmap was aligned to the healthy controls average Gradient I via the Procrustes alignment [16]. Next, the spatial correlation between the resulting OCD maps and the control eigenmap was calculated. To assess if a normalisation of OCD individual resting state eigenmaps as a function of time linked to change in Y-BOCS scores, we compared the number of OCD subjects showing greater eigenmap similarity with controls at follow-up relative to baseline. Specifically, we tested for a difference in the association with control eigenmaps in three OCD groups: (i) OCD subjects who at follow up showed no improvement or increased symptoms compared to baseline; (ii) subjects who showed small symptoms’ improvement (change in Y-BOCS >0% but <35%), and (iii) subjects who experienced a large improvement in symptoms (change in Y-BOCS $\geq$35%). The criteria to differentiate group (ii) from group (iii) was in line with [17].

We performed a confirmatory (sensitivity) analysis to assess if the eigenmap normalisation linked to a reduction in OCD symptoms replicated once noisy individual maps were removed. To exclude individual eigenmaps in an unbiased fashion, we used a leave-one-out (LOO) procedure. In the control group, we calculated individual eigenmaps and correlated each eigenmap to those calculated from the average similarity matrix of the remaining *n-1* control subjects after Procrustes alignment [16]. We then looked for factors associated with a poor correlation to the group average, including the spatial density of the minimally connected graph and the variance explained by the first gradient. Cut-offs identified within the control LOO analysis were applied to the OCD subjects in order to perform the confirmatory (sensitivity) analysis (Supplementary Figure 3).

**Supplementary Figures**

Supplementary Figure 1. Schematic of the analyses.

Supplementary Figure 2. Comparison of the right striatal average eigenmap derived using HCP data (as in Tian et al., 2020) and the comparable eigenmap calculated using data from the control group of this study.

Supplementary Figure 3. Sensitivity analysis of the association between changes in Gradient I across time and fluctuations in OCD symptoms.

Supplementary Figure 4. Changes in resting fMRI functional gradients in controls and OCD for the left striatum.

Supplementary Figure 5. Visualisation of brain voxels showing a group difference from voxel permutation tests (Figure 2G and H).

Supplementary Figure 6. Link between changes in striatal functional topology and optimal DBS targets.

Supplementary Figure 7. Relationship between OCD-related changes in Gradient I and total Y-BOCS scores at baseline.

Supplementary Figure 8. Examples of Gradient I in OCD subjects showing a very low correlation to the average Gradient I of controls.

Supplementary Figure 9. The change in correlation to Healthy Control Gradient I against the improvement in Y-BOCS.

Supplementary Figure 10. Associations between change in Gradient I magnitude and change in total Y-BOCS scores.

Supplementary Figure 11. Second task-specific right striatum functional gradients (Gradient II) in controls and OCD.

Supplementary Figure 12. Group average similarity matrices (η^2).

Supplementary Figure 13. The variance explained by Gradients in the right striatum.

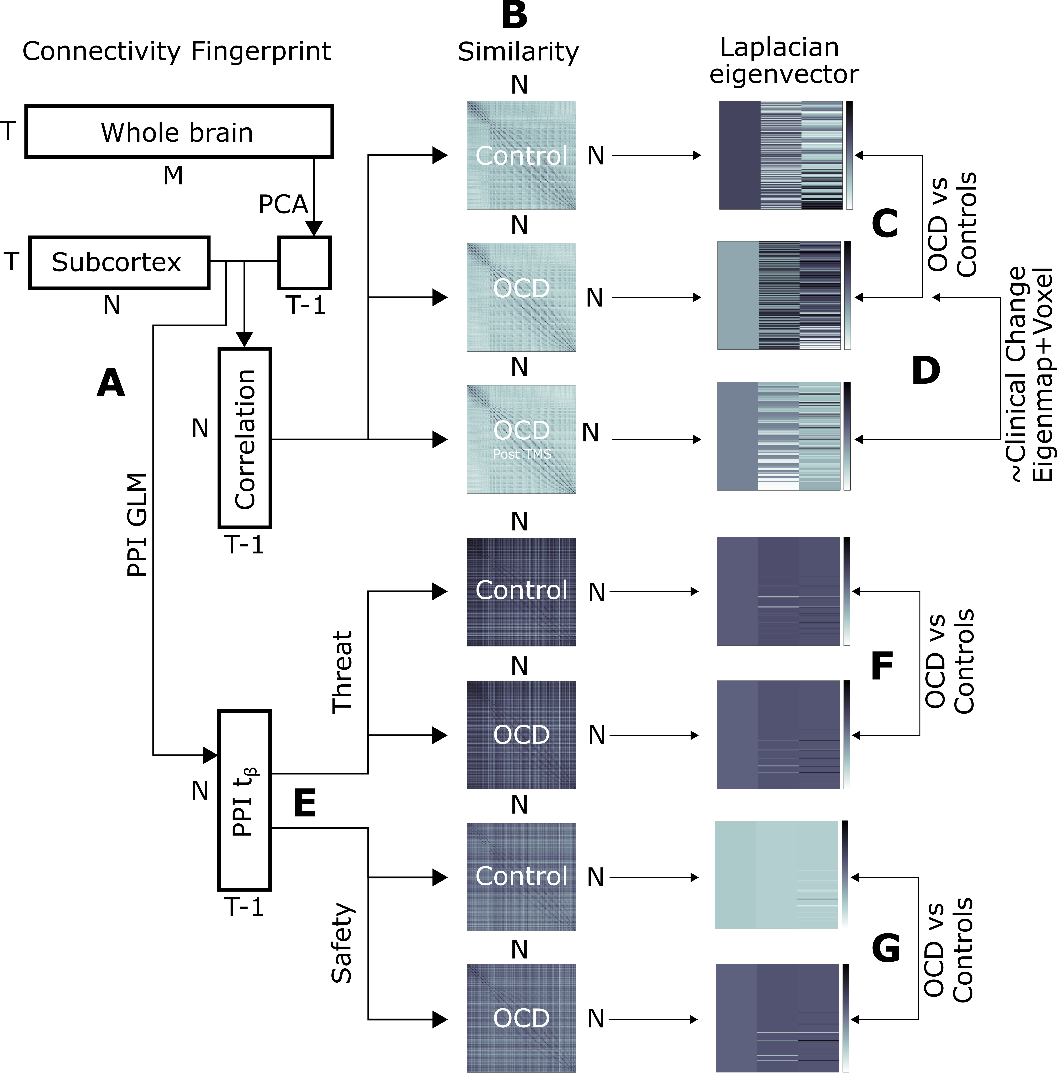

**Supplementary Figure 1.** *Schematic of the analyses.* **A)** The grey matter time series were dimensionally reduced via PCA. The resulting T-1 components were correlated to the subcortical time series. **B)** After calculating the similarity matrices, the graph Laplacian was applied to the minimally connected network (Methods). **C)** Putative differences in the eigenvector magnitude between OCD (baseline) and controls were compared via permutation testing. **D)** We quantified changes in the correlation of individual OCD eigenvectors to the average control eigenvector. Next, we assessed if the OCD eigenvectors degree of normalisation was linked with improvements in symptom severity (total Y-BOCS score). We also correlated the change in eigenvector magnitude to change in symptom severity in brain voxels that showed a group difference. **E)** A psycho-physiological interaction (PPI) was applied to isolate task-based connectivity. The t statistics linked to the PPI $\beta$coefficients relating to the threat and safety conditions were used to calculate similarity matrices (Methods). **F)** The eigenvector magnitudes in the Threat task condition were then compared between control and OCD groups. **G)** The eigenvector magnitudes in the Safety task condition were also compared between the control and OCD groups.

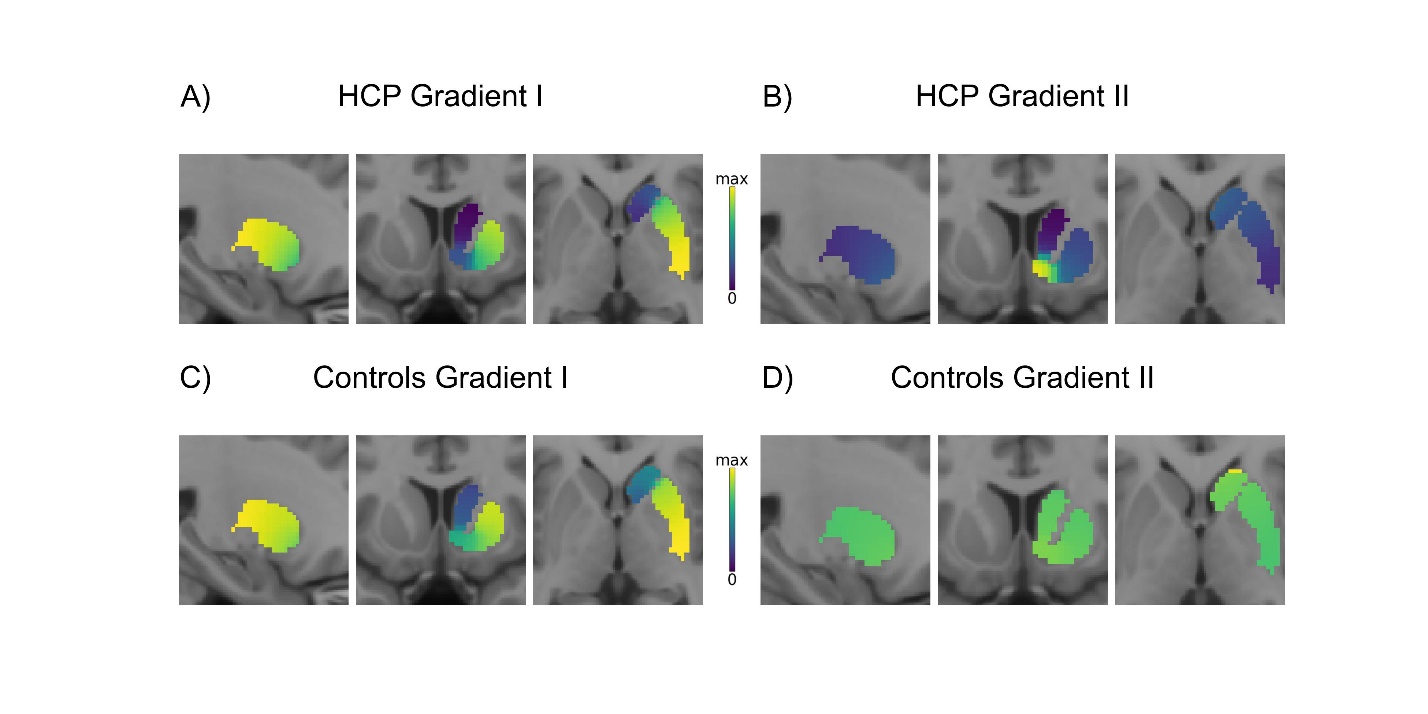

**Supplementary Figure 2.** Comparison of the right striatal average eigenmap derived using HCP data (as in Tian et al., 2020) and the comparable eigenmap calculated using data from the control group of this study. **A)** HCP Gradient I, variance explained 19.9%. **B)** HCP Gradient II, variance explained 4.9%. **C)** Control Gradient I, variance explained 20.7%. **D)** Control Gradient II, variance explained 6.0%. A similar caudate-NAcc-putamen pattern is seen in the HCP and controls Gradient I (**A** and **C**). Meanwhile, the Gradient II topology differed between the HCP and our controls (**B** and **D**). Imaging slice coordinates: x = 25, y = 10, z = 0 (MNI space).

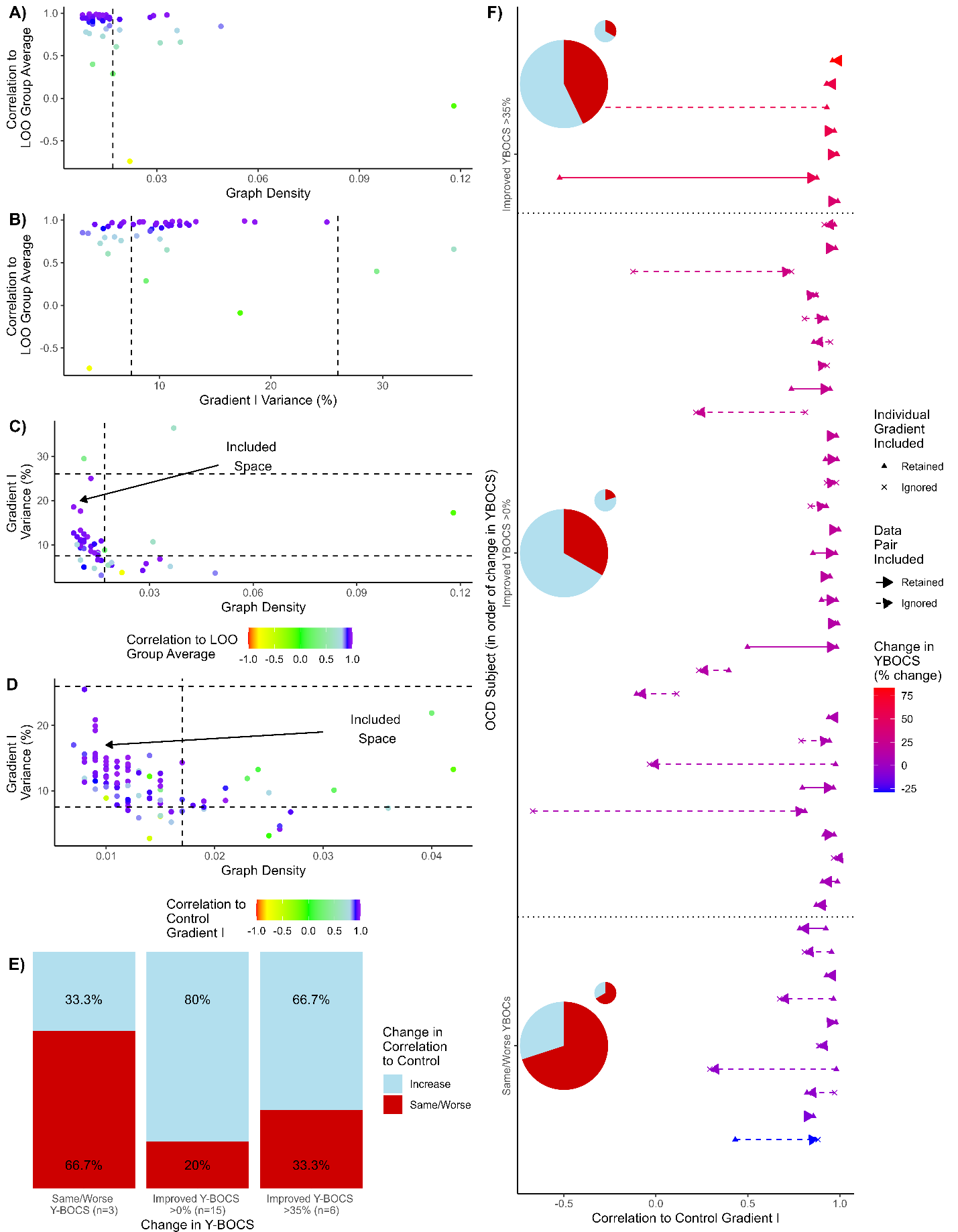

**Supplementary Figure 3.** *Sensitivity analysis of the association between changes in Gradient I across time and fluctuations in OCD symptoms.* This analysis tests the stability of our original findings by removing individuals with OCD showing Gradient I maps that may be spurious (see Methods). A Leave-One-Out (LOO) procedure was applied to establish objective cut-offs for the inclusion of individual eigenmaps in this sensitivity analysis. Individuals OCD and control resting state eigenmap were correlated to the *n-1* control group averages. Cut-offs included: **(A)** high minimum graph density (>1.7%) to obtain a fully connected graph, and **(B)** low variance explained by Gradient I (<7.5%) or high variance explained by Gradient I (>26%). Adopting these criteria removed 44% of controls **(C)** and 30% of the timepoints of individuals with OCD **(D)**. Five subjects had both individual eigenmaps falling outside the LOO criteria, nine subjects had only the baseline fall outside, and nine subjects had their post treatment eigenmap fall outside, retaining 51.0% (N=24) individuals compared to the original analyses (N=47). Results from this sensitivity analysis confirmed that in OCD individuals showing clinical improvement across time, a higher proportion showed Gradient I approached the control group gradient **(E).** While stronger, this trend remained not statistically significant (p=0.22, Fisher’s exact test). **F)** The panel shows the correlation (x-axis) between the average control Gradient I and individual OCD gradients, ordered by percent change in Y-BOCS (y-axis). Most individual gradients have a high correlation to the control Gradient I (*r*>0.8). Pie charts show the proportion of OCD individuals Gradient I that become more correlated to the Control Gradient 1 (light blue) or less correlated (red) between the first timepoint and the second time-point (Top: group showing decrease in Y-BOCS scores by over 35% at the second time-point; Middle: group that showed decrease Y-BOCS scores at the second time-point; Bottom: group showing same/worse Y-BOCS scores at time-point two;). Larger pie charts reflect results when all subjects (N=47, as Figure 3) were included in the analyses, while smaller pie charts represent findings excluding Gradients I that were considered possibly spurious (as in **E**).

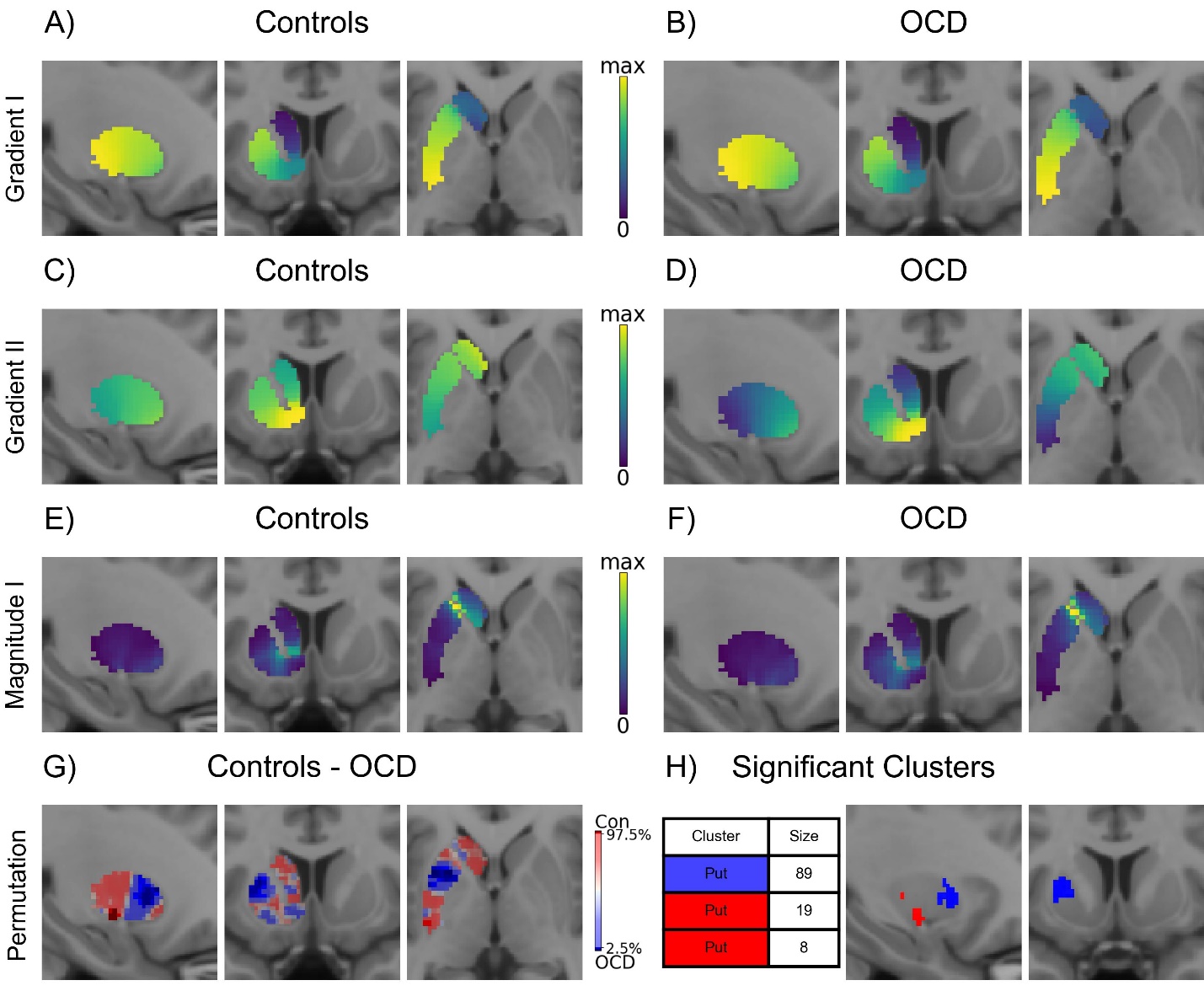

**Supplementary Figure 4.** *Changes in resting fMRI functional gradients in controls and OCD for the left striatum.* **A)** The Gradient I eigenmap for controls, which explained 15.4% of the variance. **B)** The OCD Gradient I eigenmap, which explained 17.9% of the variance. **C)** The Gradient II eigenmap for controls, which explained 3.7% of the variance. **D)** For the OCD Gradient II eigenmap the variance explained was 4.0%. **E)** The Gradient I magnitude for controls. **F)** The Gradient I magnitude for OCD participants. The maximum magnitude in Gradient I was 0.582 for both control (**E**) and OCD (**F**). Panel **G** shows the spatial extent of the percentile voxel-wise group magnitude differences (n=1000 permutations). Dark red indicates the Magnitude was higher in the Control gradient I, and Dark Blue indicate the Magnitude was higher in the OCD gradient I. Imaging slice coordinates for *A-G*: x = -25, y = 10, z = 0. **H)** Clusters of voxels with significantly different magnitudes (cluster-extent p<0.05), with all three located in the putamen. Imaging slices in H: x = -28, y = 8 (MNI space).

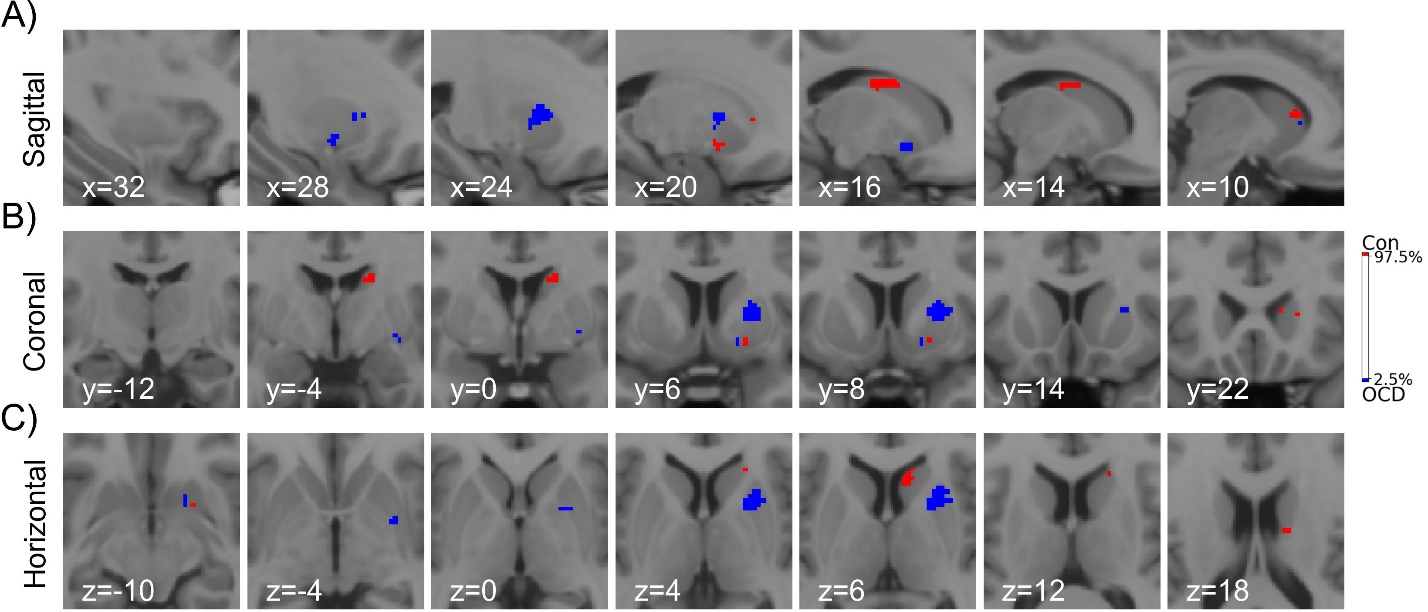

**Supplementary Figure 5.** *Visualisation of brain voxels showing a group difference from voxel permutation tests* (Figure 2G and H): **(A)** sagittal, **(B)** coronal, and **(C)** horizontal slices. Voxels coloured red indicate a higher average magnitude in controls Gradient I, while blue colour indicates a higher magnitude in OCD. All voxels surviving the voxel level group permutations (p<0.05, uncorrected for multiple tests) are shown. Note that not all voxels survive the cluster permutation test.

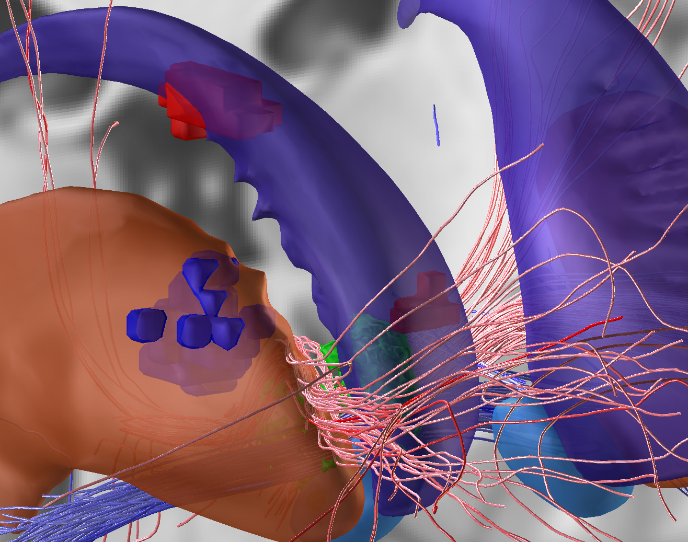

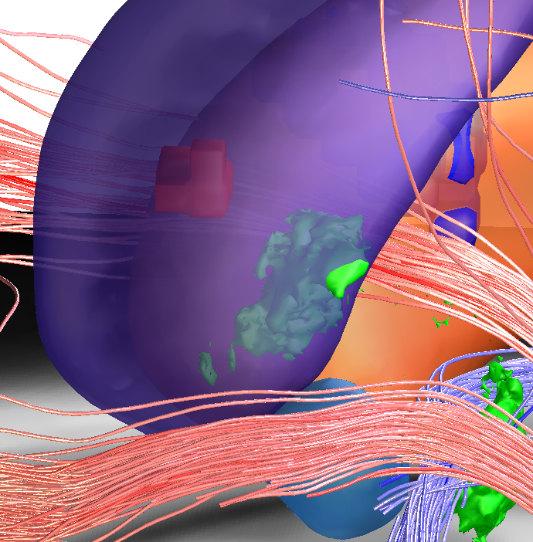

**Supplementary Figure 6.** *Link between changes in striatal functional topology, OCD-related white-matter tracts, and optimal DBS targets.* Striatal clusters showing statistically significant group differences in Gradient I magnitudes, the ‘sweet-spot’ identified by Meyer, Hollunder [18] and the OCD response tract from Li, Baldermann [19]. Clusters that showed a significant group difference (Figure 2H) are shown: blue colour indicates a higher Gradient I magnitude in OCD than controls, while red indicates a lower Gradient I magnitude in OCD than controls. The significant clusters in our results do not overlap with, nor are immediately adjacent to, the DBS response sweet spots nor OCD response tract (pink). Though, the dorsal sweet spot (shown in green) [18] is located near the mid-anterior caudate cluster of voxels with lower Gradient I magnitude in OCD. Further, the OCD response tract runs between the putamen and caudate, in close proximity to the cluster of voxels with higher Gradient I magnitude in the putamen (left). Images made in Lead Anatomy using the OCD Tract Target Atlas [19]. Note that the anterior caudate cluster (red, right) is at the anterior surface of the caudate in the Melbourne Subcortical atlas [8], though appears inside the caudate in this atlas.

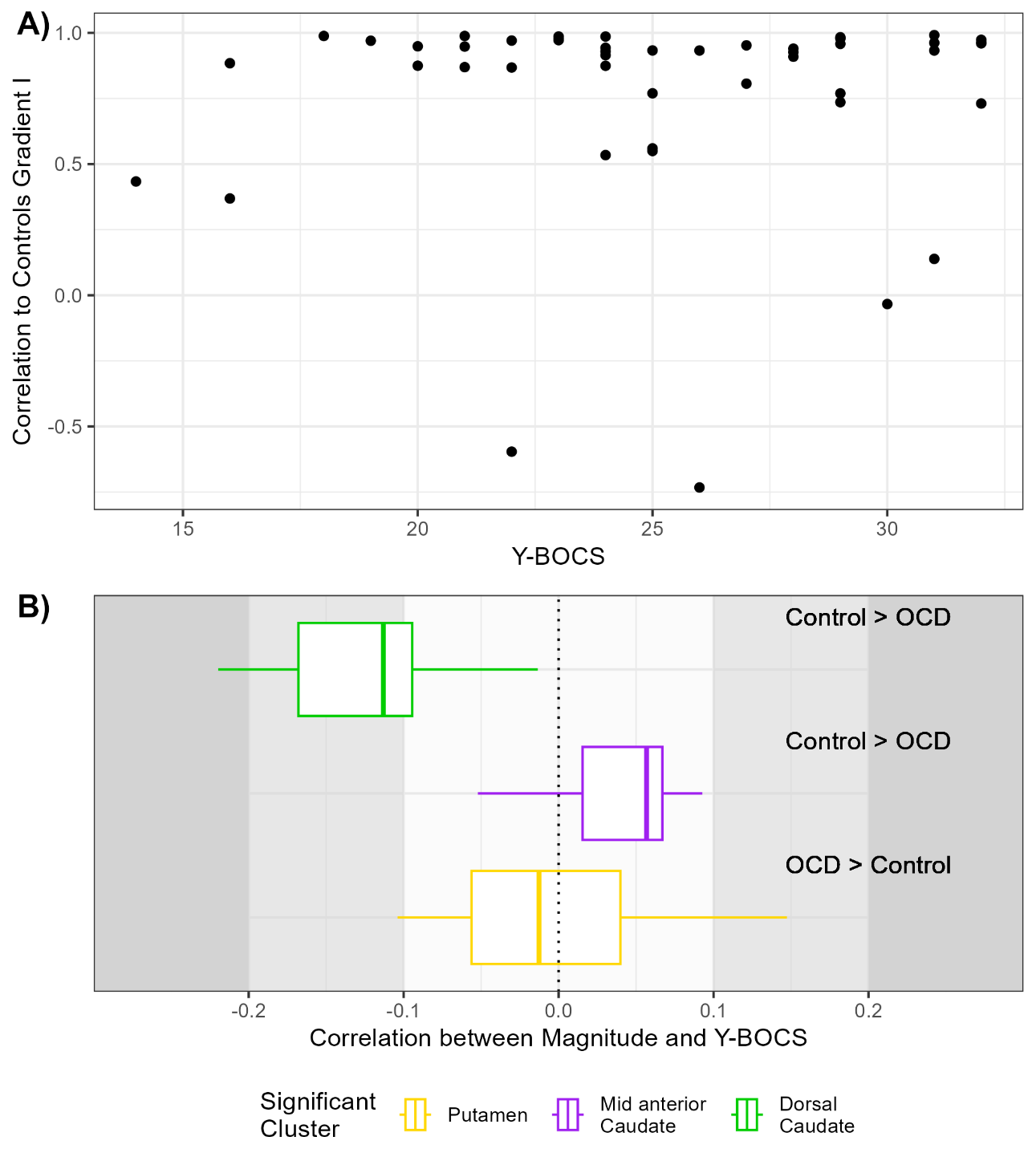

**Supplementary Figure 7**. *Relationship between OCD-related changes in Gradient I and total Y-BOCS scores at baseline*. The top panel **(A)** presents a continuous assessment of the tested association (Spearman r=0.025). **B)** The panel shows distributions of the relationship between baseline Y-BOCS scores and the Gradient I magnitude in the voxels defining clusters showing a significant group effect (see Figure 2G and H).

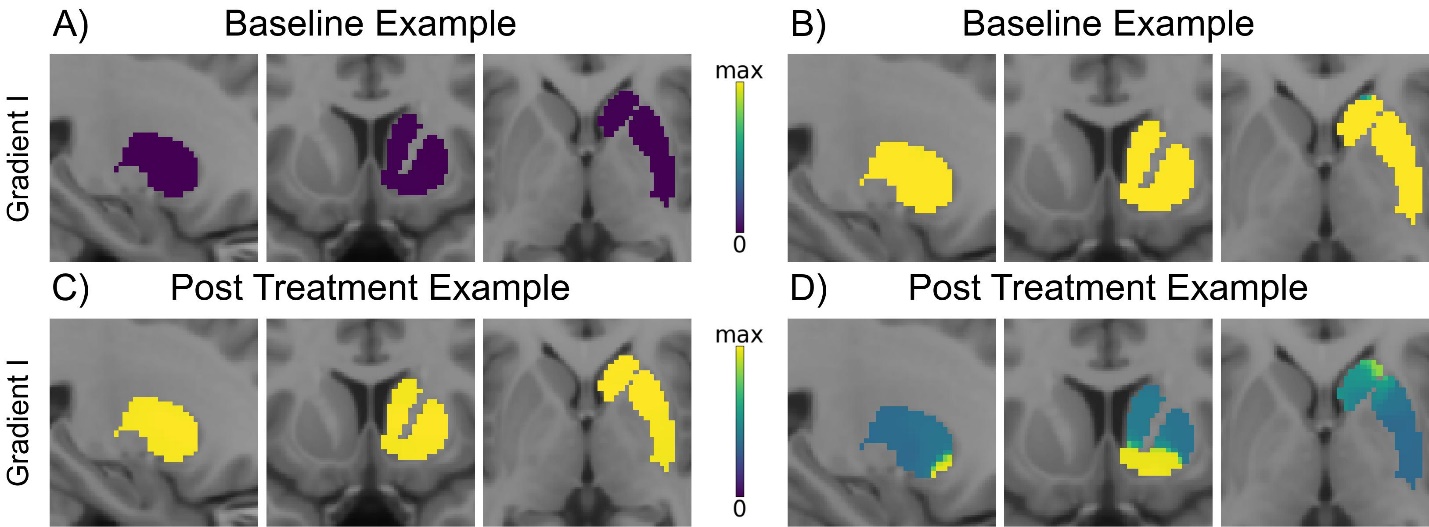

**Supplementary Figure 8.** *Examples of Gradient I in OCD subjects showing a very low correlation to the average Gradient I of controls.* **A)** The panel shows a Gradient I from an individual with OCD at the baseline timepoint with a very low correlation (r=-0.12) to the average control group Gradient I. This individual Gradient I explained 13.3% of the variance in the data, and was calculated on a map thresholded to 4.2% of the density (criteria failed, see Supplementary Figure 3). There is no evidence of the caudate-NAcc-putamen pattern (as seen in Figure 2A and B). **B)** Gradient I from an individual with OCD at the pre-treatment timepoint with a very low correlation (r=0.11) to the average control group Gradient I. This individual Gradient I explained 13.3% of the variance in the data, and was calculated on a map thresholded to 2.4% of the density (criteria failed, see Supplementary Figure 3). There is no evidence of the caudate-NAcc-putamen pattern. **C)** Gradient I from an individual with OCD at the post-treatment timepoint with a very low correlation (r=-0.04) to the average control group Gradient I. This individual Gradient I explained 3.1% of the variance in the data, and was calculated on a map thresholded to 2.5% of the density (criteria failed, see Supplementary Figure 3). There is no evidence of the caudate-NAcc-putamen pattern. **D)** Gradient I from an individual with OCD (same as Supplementary Figure 7B) at the post-treatment timepoint with a very low correlation (r=-0.11) to the average control group Gradient I. This individual Gradient I explained 12.2% of the variance in the data, and was calculated on a map thresholded to 1.4% of the density. There is no evidence of the caudate-NAcc-putamen pattern, and appears more similar to the Gradient II topology (Supplementary Figure 2B). This individual passed the LOO-identified criteria (Supplementary Figure 3), but due to the baseline not passing the LOO criteria (B) the individual was not included in the sensitivity analysis.

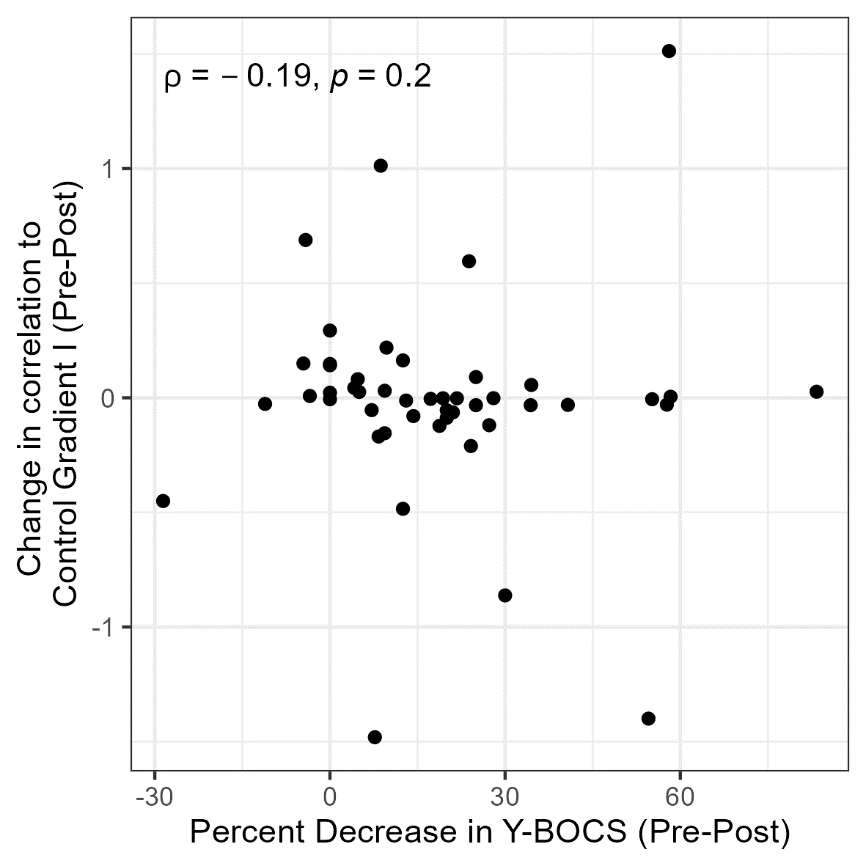

**Supplementary Figure 9.** *The change in correlation to Healthy Control Gradient I against the improvement in Y-BOCS.* When considered as continuous measures, the relationship between the change in correlation to Control Gradient I and the improvement in OCD symptoms (decrease in Y-BOCS) has a weak negative relationship ($\rho=-0.19$), indicating that an increase in correlation to the Control Gradient I is linked to a decrease in symptoms. As shown in Supplementary Figure 3, there are many outliers in this relationship, caused by poor individual fits.

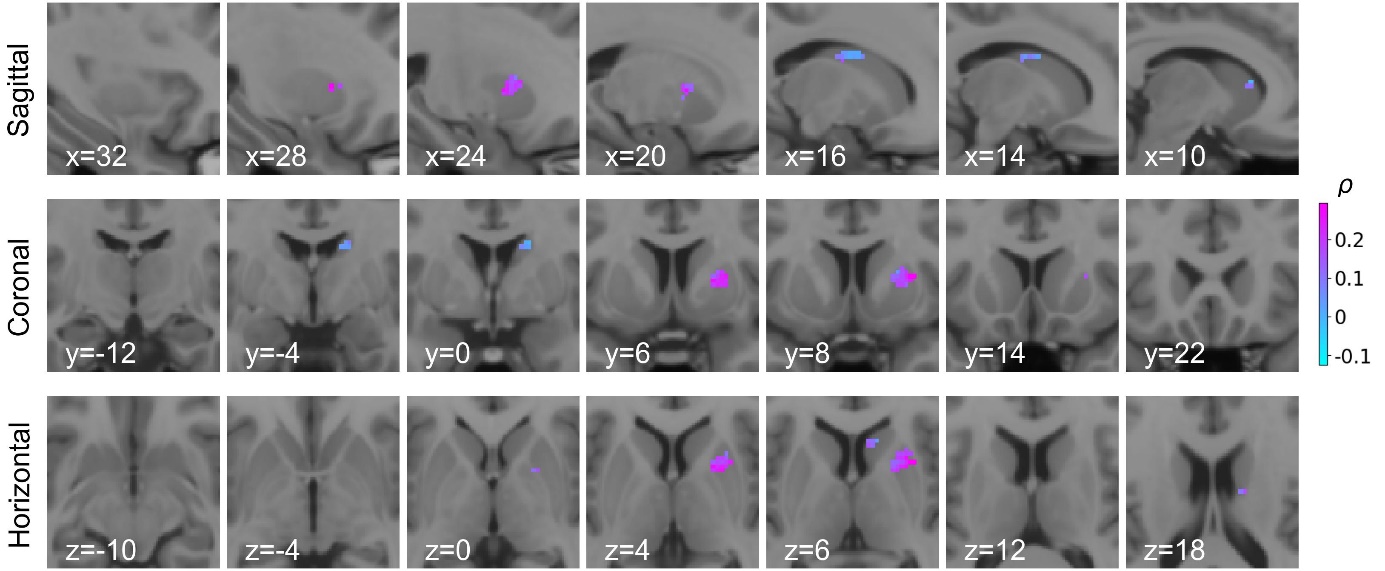

**Supplementary Figure 10.** *Associations between change in Gradient I magnitude and change in total Y-BOCS scores*. The figure depicts the associations (spearman correlation, $\rho$) between voxel-level change in Gradient I magnitude and total Y-BOCS scores (Figure 2D) in clusters showing a significant group effect (Figure 1G and H). Voxels coloured pink indicate a stronger positive association between improvement in OCD symptoms and changes in Gradient I magnitude toward the Control Gradient I. The large cluster of significant voxels in the putamen (Supplementary Figure 5) contains the voxels with the strongest associations with improvement.

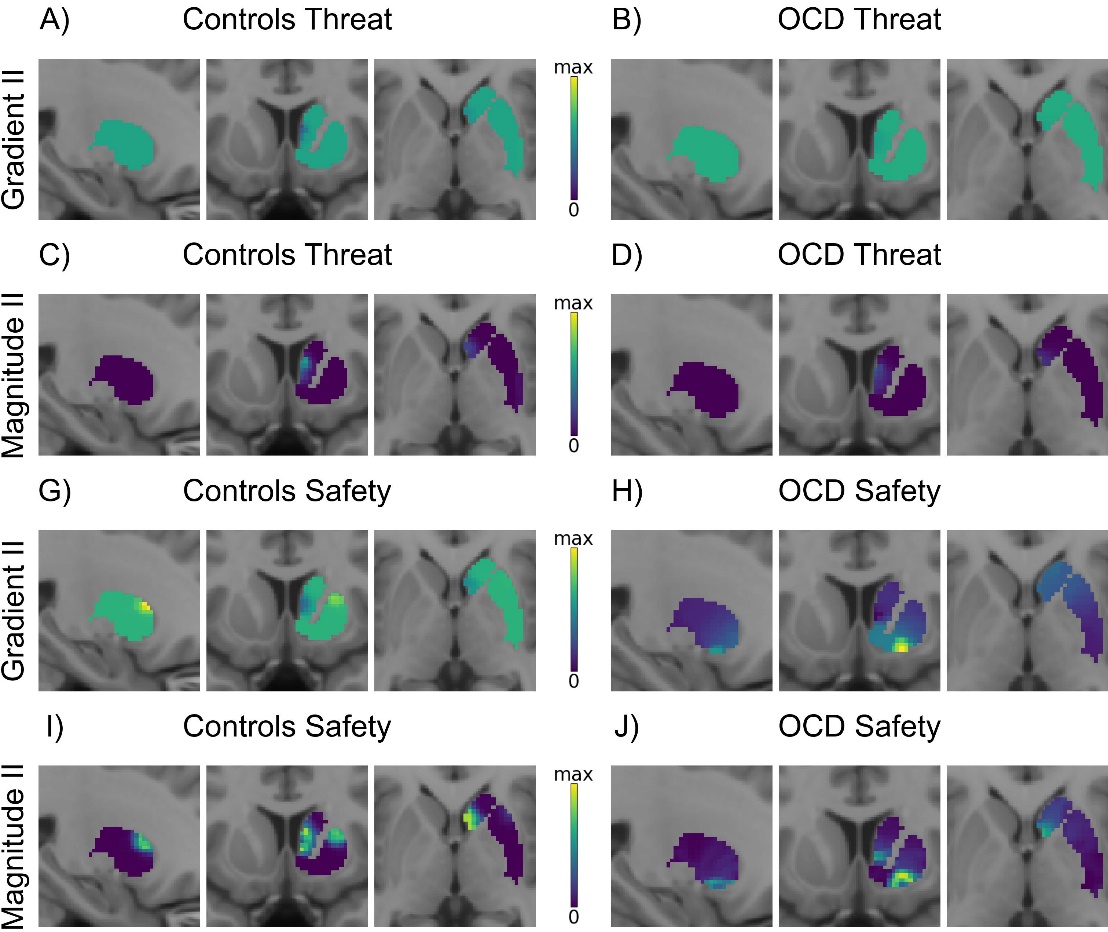

**Supplementary Figure 11.** Second task-specific right striatum functional gradients (Gradient II) in controls and OCD. **A)** The control group Gradient II eigenmap for the threat condition. This gradient explained 8.2% of the variance. **B)** This panel shows the OCD Gradient II eigenmap for the threat-reversal condition. In OCD, this gradient explained 6.7% of the variance. **C)** Average gradient II magnitude for the threat-reversal condition in controls. The maximum magnitude was 4.822. **D)** Average gradient II magnitude of the threat condition in OCD. The maximum magnitude was 3.098. **G)** Representation of the control group average Gradient II eigenmap associated with the safety-reversal task condition. The variance explained was 2.5%. **H)** The average Gradient II eigenmap for the safety-reversal condition for OCD. In OCD, this gradient only explains 3.8% of the variance. **I)** The average Gradient II magnitude for the safety-reversal condition in controls. The maximum magnitude was 2.255. **J)** The safety-reversal average Gradient II magnitude in OCD, showing a maximum magnitude of 2.019. Imaging slice coordinates: x = 25, y = 10, z = 0.

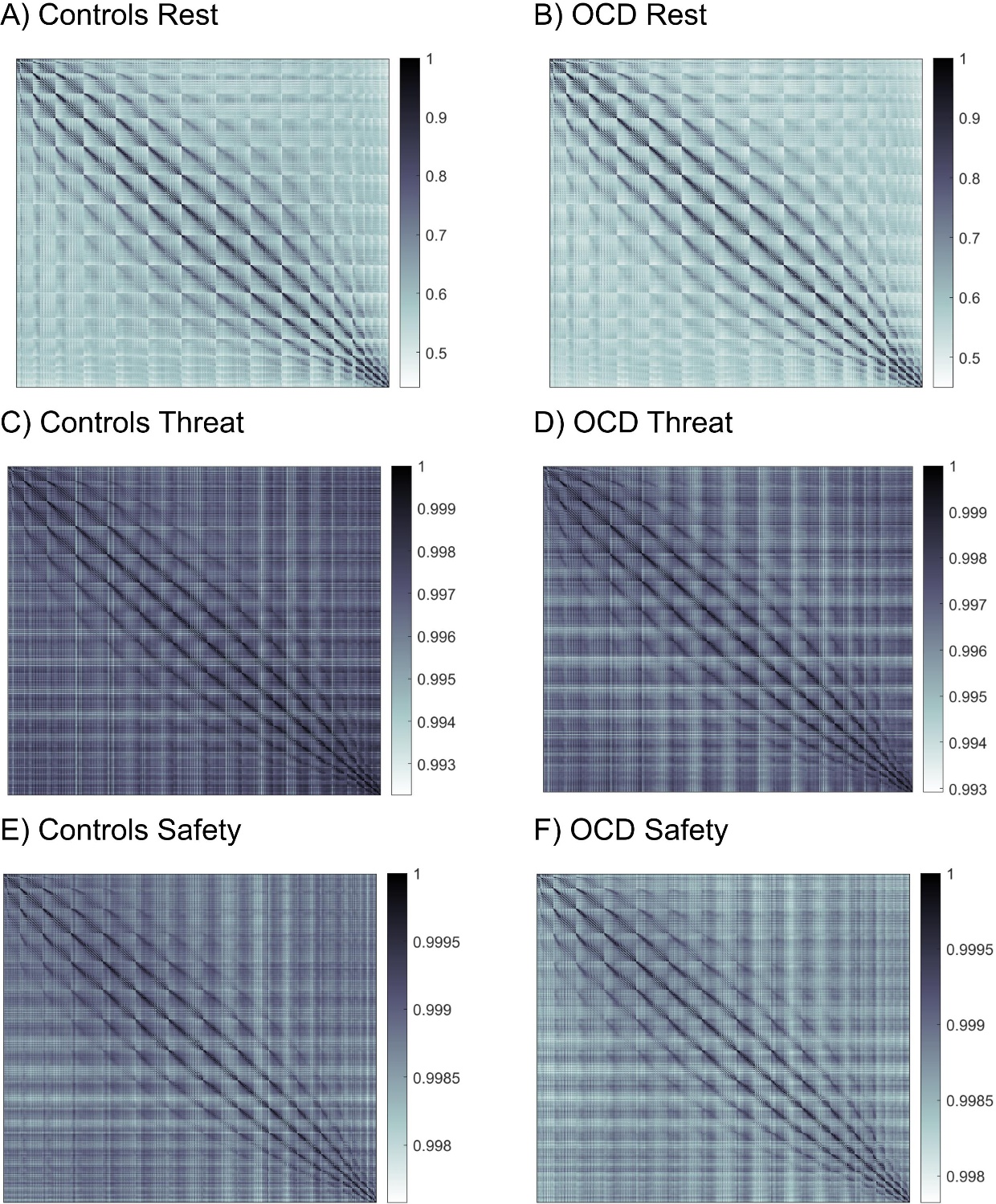

**Supplementary Figure 12.** *Group average similarity matrices (η^2)*. **A)** The Controls resting state similarity matrix for the striatum voxels. Similarity ranges from 0.440 to 1, with a mean (SD) similarity of 0.603 (0.071). **B)** The OCD resting state similarity matrix for the striatum voxels. Similarity ranges from 0.449 to 1, with a mean (SD) similarity of 0.591 (0.070). **C)** The Controls threat reversal state similarity matrix for the striatum voxels. Similarity ranges from 0.992 to 1, with a mean (SD) similarity of 0.997 (8.74×10^-4). **D)** The OCD threat reversal state similarity matrix for the striatum voxels. Similarity ranges from 0.993 to 1, with a mean (SD) similarity of 0.997 (8.01×10^-4). **E)** The Controls safety reversal state similarity matrix for the striatum voxels. Similarity ranges from 0.998 to 1, with a mean (SD) similarity of 0.999 (2.42×10^-4). **F)** The OCD safety reversal state similarity matrix for the striatum voxels. Similarity ranges from 0.998 to 1, with a mean (SD) similarity of 0.999 (2.36×10^-4).

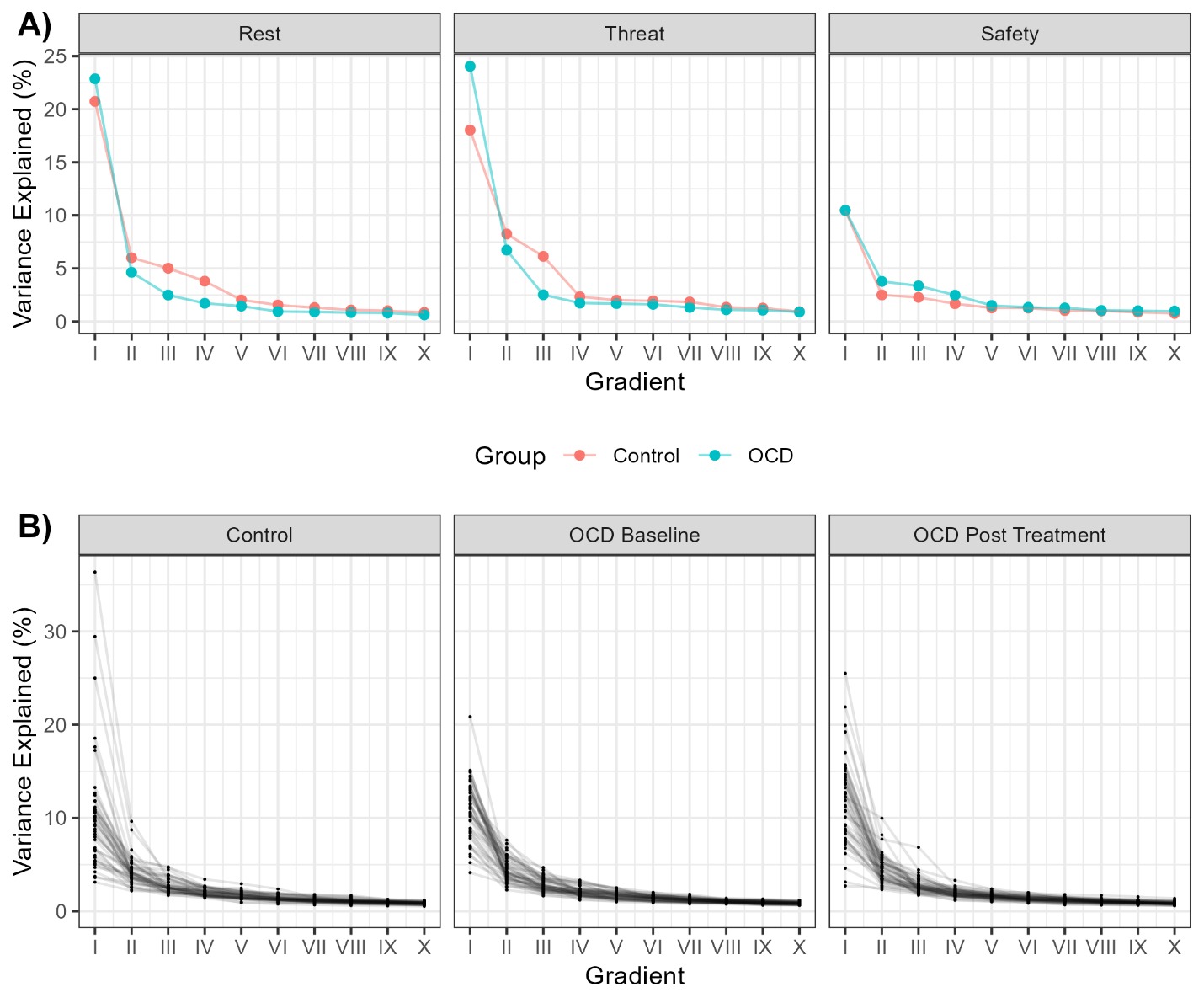

**Supplementary Figure 13.** *The variance explained by Gradients in the right striatum.* **A)** The percentage variance explained in the group average gradients for the controls and the OCD participants at rest, and in each of the threat-safety reversal conditions. **B)** The percentage variance explained in the individual gradients for the controls, and the OCD participants at both timepoints.

**Supplementary Material References**

1.	Naze, S., et al., *Mechanisms of imbalanced frontostriatal functional connectivity in obsessive-compulsive disorder.* Brain, 2022. **146**(4): p. 1322-1327.

2.	Hearne, L.J., et al., *Revisiting deficits in threat and safety appraisal in obsessive-compulsive disorder.* Human Brain Mapping, 2023. **44**(18): p. 6418-6428.

3.	Cocchi, L., et al., *Effects of transcranial magnetic stimulation of the rostromedial prefrontal cortex in obsessive–compulsive disorder: a randomized clinical trial.* Nature Mental Health, 2023. **1**(8): p. 555-563.

4.	Goodman, W.K., et al., *The Yale-Brown Obsessive Compulsive Scale: II. Validity.* Archives of General Psychiatry, 1989. **46**(11): p. 1012-1016.

5.	Esteban, O., et al., *fMRIPrep: a robust preprocessing pipeline for functional MRI.* Nature Methods, 2019. **16**(1): p. 111-116.

6.	Abraham, A., et al., *Machine learning for neuroimaging with scikit-learn.* Frontiers in Neuroinformatics, 2014. **8**.

7.	Wang, H.-T., et al., *Continuous evaluation of denoising strategies in resting-state fMRI connectivity using fMRIPrep and Nilearn.* PLOS Computational Biology, 2024. **20**(3): p. e1011942.

8.	Tian, Y., et al., *Topographic organization of the human subcortex unveiled with functional connectivity gradients.* Nature Neuroscience, 2020. **23**(11): p. 1421-1432.

9.	Borne, L., et al., *Functional re-organization of hippocampal-cortical gradients during naturalistic memory processes.* Neuroimage, 2023. **271**: p. 119996.

10.	Glasser, M.F., et al., *Using temporal ICA to selectively remove global noise while preserving global signal in functional MRI data.* Neuroimage, 2018. **181**: p. 692-717.

11.	Masharipov, R., et al., *Comparison of whole-brain task-modulated functional connectivity methods for fMRI task connectomics.* Communications Biology, 2024. **7**(1): p. 1402.

12.	Gaudes, C.C., et al., *Detection and characterization of single-trial fMRI bold responses: Paradigm free mapping.* Human Brain Mapping, 2011. **32**(9): p. 1400-1418.

13.	Friston, K.J., et al., *Psychophysiological and modulatory interactions in neuroimaging.* Neuroimage, 1997. **6**(3): p. 218-29.

14.	McLaren, D.G., et al., *A generalized form of context-dependent psychophysiological interactions (gPPI): a comparison to standard approaches.* Neuroimage, 2012. **61**(4): p. 1277-86.

15.	Lindquist, M.A. and A. Mejia, *Zen and the art of multiple comparisons.* Psychosom Med, 2015. **77**(2): p. 114-25.

16.	Vos de Wael, R., et al., *BrainSpace: a toolbox for the analysis of macroscale gradients in neuroimaging and connectomics datasets.* Communications Biology, 2020. **3**(1): p. 103.

17.	Mataix-Cols, D., et al., *Operational Definitions of Treatment Response and Remission in Obsessive-Compulsive Disorder Capture Meaningful Improvements in Everyday Life.* Psychotherapy and Psychosomatics, 2022. **91**(6): p. 424-430.

18.	Meyer, G.M., et al., *Deep Brain Stimulation for Obsessive-Compulsive Disorder: Optimal Stimulation Sites.* Biological Psychiatry, 2024. **96**(2): p. 101-113.

19.	Li, N., et al., *A unified connectomic target for deep brain stimulation in obsessive-compulsive disorder.* Nature Communications, 2020. **11**(1): p. 3364.
